# Endothelial Cell-Derived Extracellular Vesicles Elicit Neutrophil Deployment from the Spleen Following Acute Myocardial Infarction

**DOI:** 10.1101/2020.12.03.20242875

**Authors:** Naveed Akbar, Adam T. Braithwaite, Emma M. Corr, Graeme J. Koelwyn, Coen van Solingen, Clément Cochain, Antoine-Emmanuel Saliba, Alastair Corbin, Daniela Pezzolla, Laurienne Edgar, Mala Gunadasa-Rohling, Abhirup Banerjee, Daan Paget, Charlotte Lee, Eleanor Hogg, Adam Costin, Raman Dhaliwal, Errin Johnson, Thomas Krausgruber, Joey Riepsaame, Genevieve E. Melling, Mayooran Shanmuganathan, Oxford Acute Myocardial Infarction Study (OxAMI), Christoph Bock, David R. F Carter, Keith M. Channon, Paul R. Riley, Irina A. Udalova, Kathryn J. Moore, Daniel Anthony, Robin P. Choudhury

**Affiliations:** Division of Cardiovascular Medicine, Radcliffe Department of Medicine, University of Oxford; NYU Cardiovascular Research Center, Department of Medicine, Division of Cardiology, School of Medicine, New York University School of Medicine; Comprehensive Heart Failure Center, University Hospital Wurzburg, Germany; Helmholtz Institute for RNA-based Infection Research (HIRI), Helmholtz-Center for Infection Research (HZI); Kennedy Institute of Rheumatology, University of Oxford; Department of Physiology, Anatomy and Genetics, University of Oxford; Sir William Dunn School of Pathology, University of Oxford; CeMM Research Center for Molecular Medicine of the Austrian Academy of Sciences, Vienna, Austria; Department of Biological and Medical Sciences, Oxford Brookes University; Acute Vascular Imaging Centre, Radcliffe Department of Medicine, University of Oxford; Department of Laboratory Medicine, Medical University of Vienna, Vienna, Austria; Department of Pharmacology, University of Oxford

## Abstract

Acute myocardial infarction rapidly increases blood neutrophils (<2 hours). Release of neutrophils from bone marrow, in response to chemokine elevation, has been considered their source, but chemokine levels peak up to 24 hours after injury, and *after* neutrophil elevation. This suggests that additional non chemokine-dependent processes may be involved. Endothelial cell (EC) activation promotes the rapid (<30 minutes) release of extracellular vesicles (EVs), which are enriched in vascular cell adhesion molecule-1 (VCAM-1) and miRNA-126, and are thus a potential mechanism for communicating with remote tissues.

Here, we show that injury to the myocardium rapidly mobilises neutrophils from the spleen to peripheral blood and induces their transcriptional activation prior to their arrival at injured tissue. Ischemic myocardium leads to the generation and release of EC-derived-EVs bearing VCAM-1. EC-EV delivery to the spleen alters inflammatory gene and chemokine protein expression, and mobilises neutrophils to peripheral blood. Using CRISPR/Cas9 genome editing we generated VCAM-1-deficient EV and showed that its deletion removed the ability of EC-EV to provoke the mobilisation of neutrophils. Furthermore, inhibition of miRNA-126 *in vivo* reduced myocardial infarction size in a mouse model. Our findings show a novel mechanism for the rapid mobilisation of neutrophils to peripheral blood from a splenic reserve, independent of classical chemokine signalling, and establish a proof of concept for functional manipulation of EV-communications through genetic alteration of parent cells.

**One Sentence Summary:** Extracellular vesicles mediate neutrophil mobilisation from the spleen following acute myocardial infarction.

## Introduction

Acute myocardial infarction (AMI) is a substantial sterile injury that leads to a rapid increase in peripheral blood neutrophils *(1-5)*. Elevated peripheral blood neutrophil number post-AMI correlates with the extent of myocardial injury, degree of cardiac dysfunction, and mortality *(1-3)*. Defective neutrophil removal enhances susceptibility to cardiac rupture *(6)* and inhibition of neutrophil recruitment in AMI reduces infarct size *(1)*, but antibody depletion of neutrophils prior to AMI increases infarct size, enhances fibrosis and lowers the number of M2 macrophages in the healing myocardium *(1, 7)*. These findings suggest a complex role for neutrophils in the contexts of myocardial ischaemic injury and repair.

The bone marrow is the primary site for granulopoiesis *(8, 9)* and has been regarded as the principal source of neutrophils that are mobilised to peripheral blood after injury *(4, 10)*. Mature neutrophils are held in large numbers in the haemopoietic cords, separated from the blood by the sinusoidal endothelium *(11)*. In the current paradigm, these cells are retained in the marrow by the interaction of CXCR4 and CXCL12 (stromal cell derived factor (SDF-1α)) *(12)* and mobilised in response to soluble factors. Intravascular injection of a range of chemotactic factors, including leukotriene B4, C5a, interleukin-8 (IL-8) *(13)*, CXCL chemokines *(12, 14)* and granulocyte-colony stimulating factor (G-CSF) *(15, 16)* can drive the rapid mobilisation of neutrophils across the sinusoidal endothelium. However, numerous strands of evidence question whether chemokines derived from injured tissues are responsible for very early neutrophil mobilisation *in vivo*. Intra-cardiac mRNA levels for cytokines peak 12 hours after injury *(17)* and pro-inflammatory proteins are very modestly elevated in coronary sinus following reperfusion therapy in AMI *(18, 19)*. Furthermore, *in vivo* blood chemokine profiles peak 24 hours post-AMI and do not precede the rise in blood neutrophil counts in humans or mice, which occurs within 2 hours in mice following injury *(1, 20)*. Moreover, a putative source of chemokine generation in the acutely ischemic myocardium prior to neutrophil infiltration has not been identified.

These observations suggest that neutrophils may be mobilised from alternative reserves following injury and by mechanisms that are not dependent on chemokines. One possible source is extramedullary haematopoiesis in the spleen *(21)* from where neutrophils are mobilised to peripheral blood following bacterial infection *(22)*. By analogy, it is known that monocytes are deployed from a splenic reserve and not from the bone marrow following sterile injury in mice *(23)*, and that this can be driven by extracellular vesicles (EVs) that are derived from the vascular endothelium *(24)*.

EVs are membrane-enclosed envelopes *(25)* that are actively secreted by many cell types *(26-28)*. These vesicles bear bioactive cargo that includes proteins and microRNAs (miRNAs), which are derived from the parent cell. EV can alter the biological function and cellular status of cells locally *(29)* and remotely following liberation into the blood *(30)*. Endothelial cell (EC) derived-EVs bearing vascular cell adhesion molecule-1 (VCAM-1) are elevated in the blood following AMI *(24, 29)* and have a role in the mobilisation and transcriptional programming of splenic monocytes in AMI *(24)*.

Here, we sought to establish whether EC-EVs contribute to the mobilisation and programming of neutrophils and, if so, through which of their component parts. We hypothesised that EC-EVs released during ischaemia would localise to neutrophils in remote reserves in a process mediated by VCAM-1, which has been shown to bind to neutrophils via surface integrins *(31)*. Furthermore, we reasoned that once localised to neutrophils in reserve pools, EC-EV miRNA cargo could induce functionally relevant transcriptional programmes in those target tissues and cells prior to recruitment to the injured myocardium. An understanding of these mechanisms would immediately suggest possibilities for cell-selective immuno-modulatory interventions that are relevant in AMI and, potentially, other pathologies with an inflammatory component.

Accordingly, we investigated changes in the number and composition of EC-EVs in mice and humans in the context of AMI, examined their tissue distribution and effects on cell mobilisation and programming *in vitro* and *in vivo*. We used bioinformatics techniques to identify plausible bioactive candidates and evaluated their functional significance experimentally *in vivo* using antagomir and gene editing approaches.

## Results

### Plasma neutrophil number correlates with the extent of AMI

During acute ST-segment-elevation AMI (STEMI) in human patients (median time from onset of symptoms 3 hours) peripheral blood neutrophil number at the time of presentation correlated with the extent of ischaemic injury, as determined by oedema estimation on T2-weighted magnetic resonance imaging (MRI) images obtained within 48 hours of AMI (R^2^ =0.365, P=0.017) (**Figure 1A**) and with final infarct size, determined by late gadolinium enhancement (LGE) MRI 6 months post-AMI (R^2^=0.507, P=0.003) (**Figure 1B**).

**Figure 1.**
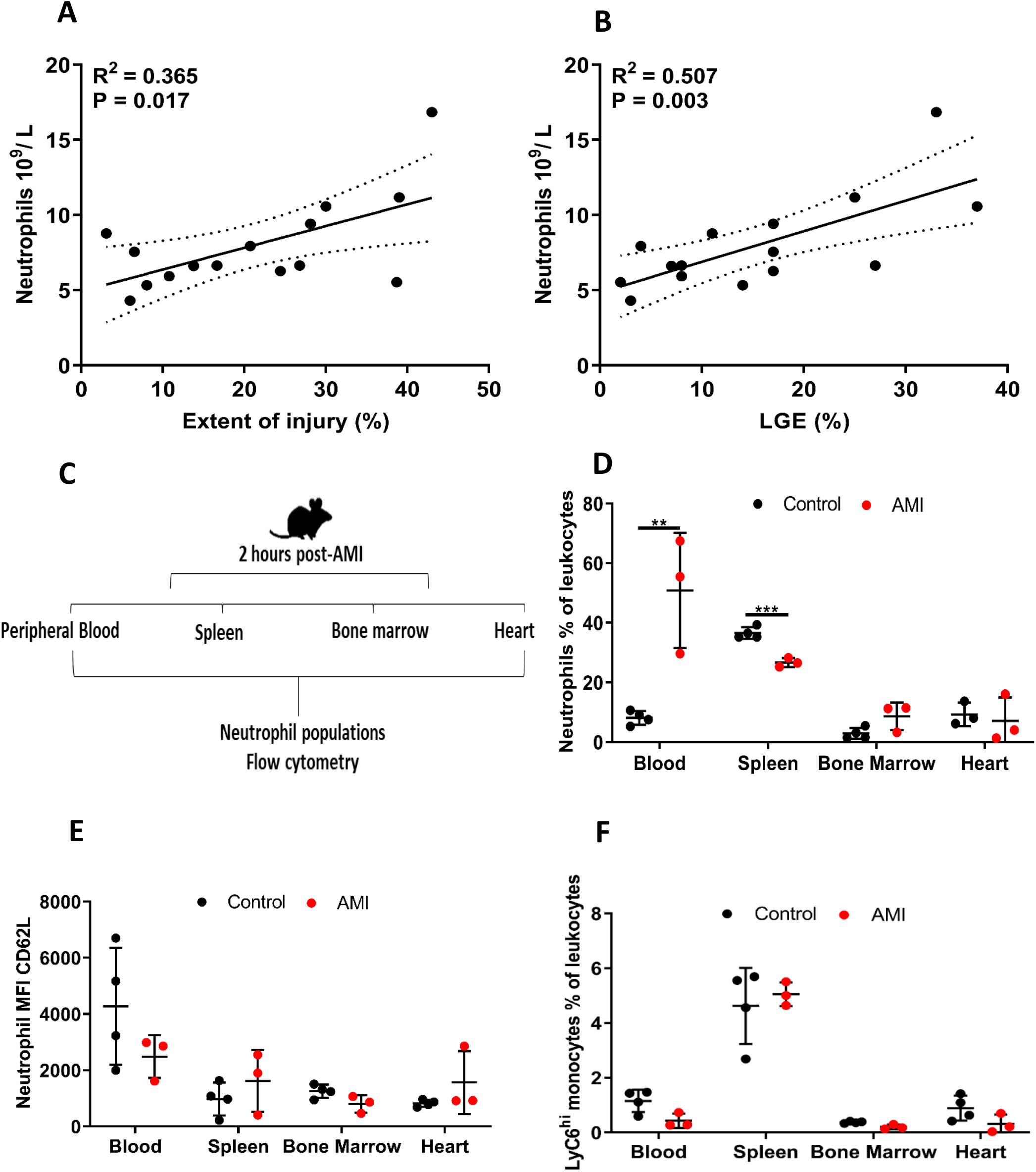
Human peripheral blood neutrophils correlate with the extent of myocardial injury in AMI. **A** Pearson’s correlation of peripheral blood neutrophil number (10 ^9^ / L) in patients experiencing AMI significantly correlated with the extent of myocardial injury (T2-weight MRI) and **B** LGE MRI 6-months post-AMI *(n* = 15). **C** Schematic representation of mouse AMI and tissue harvesting for flow cytometry. **D** Percentage of neutrophils in peripheral blood, spleen, bone marrow and heart 2 hours after AMI relative to the levels of intact controls (controls *n* = 4, AMI *n* = 3). **E** Mean fluorescence intensity (MFI) of CD62L/L-selectin on neutrophils in peripheral blood, spleen, bone marrow and heart 2 hours after AMI relative to the levels of intact controls (controls *n* = 4, AMI *n* = 3). **F** Percentage of monocytes in peripheral blood, spleen, bone marrow and heart 2 hours after AMI relative to the levels of intact controls (controls *n* = 4, AMI *n* = 3). Pearson’s correlation was used in **A** and **B**, dotted lines represent 95% confidence interval and an unpaired t test was used in **D, E** and **F** for statistical analysis. Error bars represent meanLJ±LJSD \*\**p* □< □0.01, \*\*\**p* □< □0.001.

### AMI mobilises neutrophils from the spleen

This rapid increase in peripheral blood neutrophils would be consistent with mobilisation from an existing reserve. To determine the source of neutrophil mobilisation in the immediate hours post-AMI we performed left anterior descending (LAD) artery ligation in a mouse model of AMI and analysed cell populations from multiple tissue sources, 2 hours after AMI, by flow cytometry (**Figure 1C**). AMI induced a 6.3-fold (P<0.01) increase in peripheral blood neutrophils (Live, CD45^+^, CD11b^+^, Ly6G^+^) and simultaneously lowered splenic-neutrophil number by 0.7-fold (P<0.001) (**Figure 1D**). As described previously, to obtain an indication of the mobilisation between reserves, we calculated a neutrophil mobilisation ratio *(24)* (peripheral blood neutrophils/ splenic [or bone marrow] neutrophils) and found an increase in splenic neutrophil-mobilisation (8.5-fold) (P<0.01), but no alteration in bone marrow neutrophil number relative to control animals. There was no alteration in CD62L/L-selectin (which is shed during neutrophil activation) in mobilized peripheral blood neutrophils (**Figure 1E)**. At this very early time point (2 hours post-AMI), we found no differences in LyC6^high^ monocyte number in the peripheral blood or spleen, indicating that neutrophils mobilise from the spleen prior to splenic-monocyte mobilisation *(23)* **(Figure 1F)**.

### Plasma EVs correlate with the extent of injury and neutrophil count in AMI

Following AMI EC-EVs are significantly augmented in peripheral blood, with plasma-EVs showing enrichment for miRNA-126 and the integrin VCAM-1 *(24, 29)*. In agreement with our previous findings, patients with AMI had significantly more plasma EVs at time of presentation (24.3 × 10 ^8^ ± 25.7 EV / mL) versus a 6 month follow-up measurement (11.0 × 10 ^8^ ± 12.5 EV / mL, P<0.01) but they exhibited a similar EV size distribution profile, with a significant elevation in EVs in the size range 100-200 nm diameter (**Figure 2A/B**). Plasma EV number at presentation was significantly correlated with the extent of ischaemic injury as determined by T2-weighted MRI (R^2^=0.459, P=0.006) (**Figure 2C**) and with the extent of myocardial scarring as determined by LGE at 6 months post-AMI (R^2^=0.423, P=0.009) (**Figure 2D**). We also found a highly significant relationship between plasma-EV number and peripheral blood neutrophil number (R^2^=0.753, P<0.001) at time of presentation (**Figure 2E**), consistent with a possible role for plasma-EVs in neutrophil mobilisation post-AMI.

**Figure 2.**
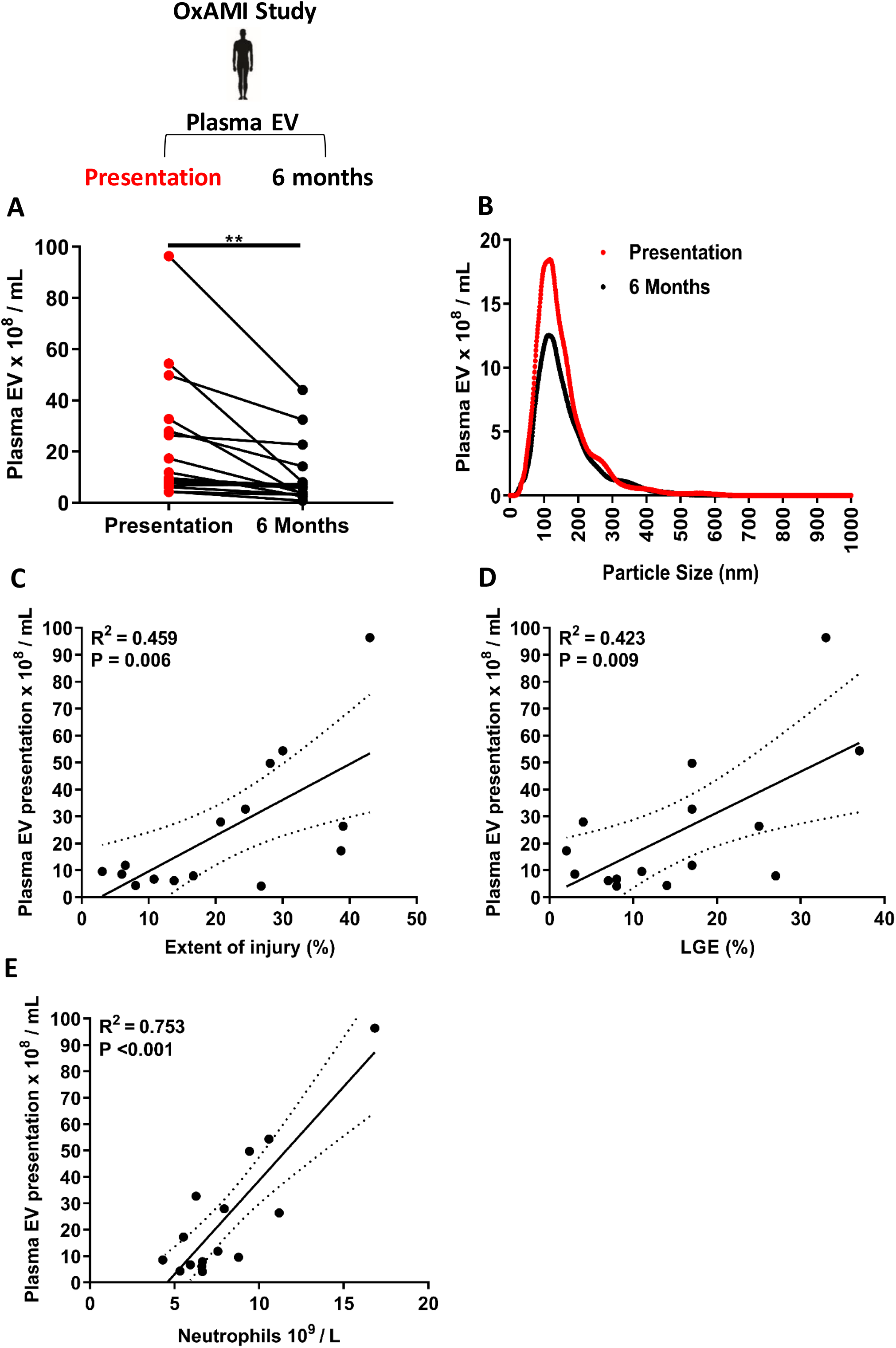
Plasma extracellular vesicles (EVs) are elevated in peripheral blood following AMI. **A** Plasma EV number (10 ^8^ / mL) at time of presentation following AMI and 6 months later in the same patients *(n* = 15). **B** Size and concentration profile of plasma EVs at time of presentation following AMI and 6 months later in the same patients *(n* = 15). Pearson’s correlation of plasma EVs at time of presentation versus: **C** extent of myocardial injury as determined by T2-weighted MRI, **D** LGE 6-months post-AMI, **E** number of peripheral blood neutrophils following AMI (10^9^ / L) *(n* = 15). A paired t test was used for statistical analysis in **A** and **B**. Pearson’s correlation was used in **C, D** and **E**, dotted lines represent 95% confidence interval for statistical analysis. \*\**p* □< □0.01

### EC-EVs are enriched for miRNA-126

In AMI, the total EV pool is enriched for EVs bearing VCAM-1 *(24)*. Endothelial cells are activated in AMI *(32)* and express VCAM-1 in response to ischaemia *in vivo (33)* and to pro-inflammatory cytokines such as tumour necrosis factor-α (TNF-α) *in vitro (34)*. In order to probe the function of EVs-derived exclusively from activated endothelial cells we studied primary human umbilical cord vein endothelial cells *in vitro*. Compared with basal conditions, treatment of endothelial cells with pro-inflammatory TNF-α activated endothelial cells as evidenced by enhanced VCAM-1 protein expression (P<0.001) (Figure 3 A) and increased EV production (P<0.001) (**Figure 3B/C)**, with a significant increase in small EVs, of similar size range (100-200 nm) to those found to increase in patients with AMI. EC-EVs displayed typical EV morphology (**Figure 3D/E)**, the EV-protein marker CD9 **(Figure 3F)** and were positive for endothelial nitric oxide synthase (eNOS). EV-depleted cell culture supernatants and cell culture media that had not been exposed to cells were negative for CD9 and eNOS, confirming robust EC-EV generation and isolation. EC-EV preparations were negative for histone and mitochondrial markers of cellular contamination, which were readily detected in endothelial cell pellets but not in isolated EC-EVs or in EV-depleted cell culture supernatants (**Figure 3F)**.

**Figure 3.**
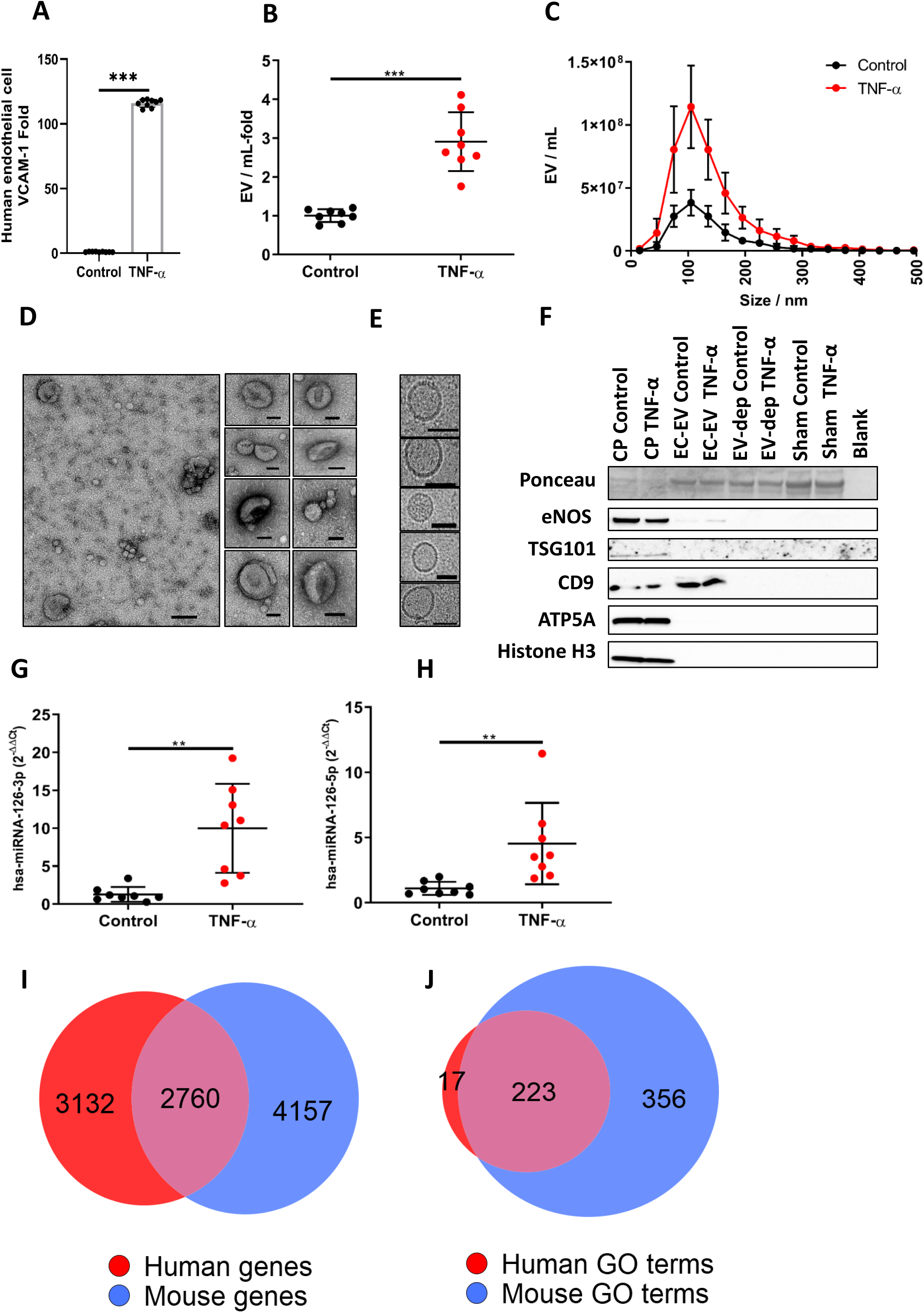
Human umbilical cord vein endothelial cells (HUVEC) release more extracellular vesicles (EVs) after inflammatory stimulation. **A** HUVECs: express more VCAM-1 following treatment with recombinant human tumour necrosis factor-α (TNF-α) (10 ng / mL) *(n* = 9 per group); **B** release more EVs *(n* = 8 per group). **C** Size and concentration profile of HUVEC derived EVs under basal conditions and after inflammatory stimulation with recombinant human TNF-α *(n* = 8 per group). **D** Transmission electron micrograph (TEM) of HUVEC-derived EVs (scale bar 100 nm) and **E** cryo-TEM HUVEC-derived EVs (scale bar 50 nm). **F** Ponceau stain and western blot of HUVEC derived EV from basal and after inflammatory stimulation with TNF-α for eNOS, TSG101, CD9, ATP5A and Histone H3. HUVEC cell pellets (CP), EV-depleted cell culture supernatants (EV-dep) and cell culture media that was not exposed to cells (sham) were used as controls. EC-EV miRNA levels of **G** hsa-miRNA-126-3p and **H** hsa-miRNA-126-5p under basal conditions and after inflammatory stimulation with TNF-α *(n* = 8 per group). miRNA-126-mRNA targets in human and mouse and their target pathways. **I** Euler plot of miRNA-126-mRNA targets from TargetScanHuman, TargetScanMouse, miRWalk, miRDB for human and the mouse. **J** Euler plot of Gene Ontology (GO) terms for miRNA-126-mRNA targets for the human and mouse. Shape areas are approximately proportional to number of genes. An unpaired t test was used in **A, B, C, G** and **H** for statistical analysis. Error bars represent mean □± SD \*\**p* □< □0.01, \*\*\**p* □< □0.001.

EC-EVs derived from pro-inflammatory stimulations show significant enrichment for miRNA-126-3p (P<0.01) (**Figure 3G)** and miRNA-126-5p (P<0.01) (**Figure 3H)**, consistent with previous observations of changes in miRNA, measured in the unselected EV pool, following AMI *(24)*.

### miRNA-126-mRNA targets cluster selectively in neutrophil motility pathways

To explore the potential role of EC-EV-miRNA-126 we employed *in silico* bioinformatics techniques. We curated miRNA-126 putative mRNA target genes from 3 different miRNA-mRNA target prediction databases for human and the mouse. Our curated lists comprised targets from TargetScanHuman, TargetScanMouse *(35)*, miRWalk *(36)* and miRDB *(37)*.

We determined whether the mRNAs putatively regulated by miRNA-126 for the human, mouse or the overlap gene set (targeted in both the human and mouse) (**Figure 3I/3J and Supplementary Table 1/2**) were present in Gene Ontology (GO) terms for neutrophil function. miRNA-126-putative-mRNA targets were significantly overrepresented when compared by Fisher’s exact test to neutrophil pathway GO terms for neutrophil migration (GO: GO1990266) in the human (P <0.001), the mouse (P<0.001) and the overlap gene set (P<0.001) (**Supplementary Table 3**). Similarly, miRNA-126-putative-mRNA targets were significantly overrepresented when compared to the neutrophil pathway GO term for neutrophil chemotaxis (GO: GO0030593) in the human (P <0.001), the mouse (P<0.001) and the overlap gene set (P<0.001) (**Supplementary Table 3**). Whereas, other neutrophil GO terms such as neutrophil mediated killing of a fungus (GO: GO0070947), neutrophil clearance (GO: GO0097350) and regulation of neutrophil mediated cytotoxicity (GO: 0070948) were not enriched (**Supplementary Table 3**), suggesting a possible role for EC-EV-miRNA-126 in orchestrating processes related to neutrophil mobilisation post-AMI.

### AMI alters neutrophil transcriptomes

To determine whether neutrophil transcriptomes are altered post-AMI we obtained peripheral blood neutrophils from newly recruited patients presenting with STEMI (N = *3)* and non-STEMI (NSTEMI) control patients (N = *3)* at time of presentation and matched-control samples one month later. STEMI patients had a greater number of differentially expressed genes at time of presentation vs NSTEMI control patients (STEMI 933 genes vs NSTEMI 8 genes) (**Figure 4A-C**). Differentially expressed neutrophil transcriptomes in STEMI patients were determined by Gene Set Enrichment Analysis (GSEA) to be positively enriched at time of presentation for inflammatory pathways, including TNF-α signalling via NF-κB (P<0.001), inflammatory response (P<0.001) and interleukin-6 (IL-6) signalling via JAK-STAT3 (P<0.001) (**Figure 4D**).

**Figure 4.**
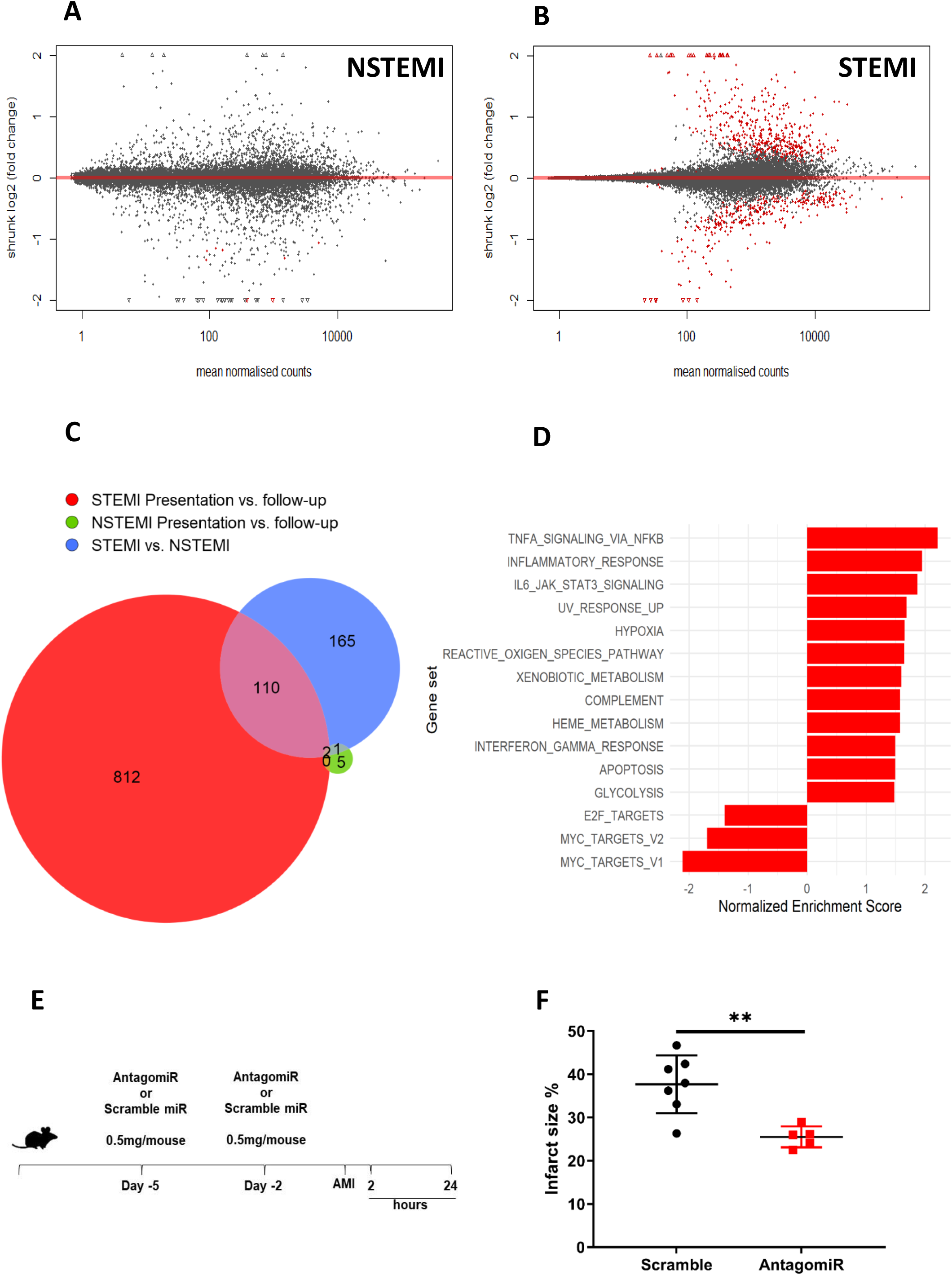
RNA sequencing of peripheral blood neutrophils following STEMI and NSTEMI patients at the time of presentation versus a control sample obtained from the same patients 1 month post-AMI *(n* = 3 per group). MA plots shows differential transcriptome at the time of presentation versus a control sample obtained from the same patients 1 month post-AMI in **A** NSTEMI and **B** STEMI patients. Significantly altered genes are highlighted in red. **C** Euler plot showing similarity and differences in the number of differentially expressed genes in NSTEMI and STEMI patients at time of presentation versus 1 month follow-control samples or between all NSTEMI and all STEMI patients *(n* = 3 per group). **D** Normalised enrichment scores are shown for significantly enriched Hallmark gene sets from gene set enrichment analysis (GSEA) of presentation versus 1 month follow-control samples from STEMI patients *(n* = 3). **E** miRNA-126 antagomiR treatment of wild-type mice prior to induction of AMI. **F** TTC staining of the myocardium 24 hours post-AMI in scramble and antagomiR treated mice (scramble *n* = 7 and antagomiR *n* = 5 per group). Significant differentially expressed (DE) genes in **A, B** and **C** were determined by adjusted *p*-values below the 5% false discovery rate (FDR) threshold. Significantly enriched gene sets in **D** were defined as having a Benjamini–Hochberg-adjusted *p*<0.05. **F** an unpaired t test was used for analysis. Error bars represent mean □± SD \*\**p* □< □0.01.

To further understand the potential target pathways for the differentially enriched genes in blood neutrophils following AMI we used GO term enrichment analysis and ranked by false discovery rate (FDR)-adjusted p-values. GO analysis showed that differentially expressed neutrophil genes at the time of presentation favoured pathways for signal recognition particle (SRP)-dependent co-translational protein targeting to membrane (GO:0006614) (P<0.001) and co-translational protein targeting to membrane (GO:0006613) (P<0.001) (**Supplementary Table 4**). Similarly, we used Reactome pathway analysis *(38)* to further explore target pathways for the differentially enrichment genes at the time of presentation and found SRP-dependent co-translational protein targeting to membrane (R-HSA-1799339) (P<0.001) and neutrophil degranulation (R-HSA-6798695) (P<0.001) to be significantly enriched (**Supplementary Table 4**).

Next, we used single cell (sc)-RNA-sequencing data to determine whether neutrophil populations in the peripheral blood of mice subjected to AMI exhibited similar transcriptomic alterations prior to recruitment to the heart. We found differential enrichment in neutrophil populations in the blood following AMI *(39)*, which favoured GO terms for neutrophil aggregation (GO:0070488) (**Supplementary Table 5 and 6**) (P<0.05), platelet activation (GO:0030168) (P<0.001) (**Supplementary Table 7**) and Reactome pathways for SRP-dependent co-translational protein targeting to membrane (R-HSA-1799339) (P<0.001) (**Supplementary Table 5 and 6)** (P<0.001) and platelet activation, signalling and aggregation (R-HSA-76002) (P<0.001) (**Supplementary Table 7**).

These analyses show that in both human and mouse neutrophils transcription is differentially regulated in AMI. We further explored commonality between the human and the mouse transcriptome response following AMI using GO terms (biological process, molecular function and cellular component) and Reactome pathways analysis. There were significant overlaps between the genes that are differentially expressed following AMI in the blood of the human and the mouse (P<0.001) (**Supplementary Table 8**) and significant similarity in target pathways between the human and mouse (**Supplementary Table 9**), demonstrating that patterns of gene expression are conserved and occur *prior* to recruitment to the injured heart. We ranked the significant comparisons by size of their intersect and found that sc-RNA-sequencing mouse blood neutrophil populations shared 38% similarity (P<0.001) and 37% similarity (P<0.001), respectively, to target pathways in human neutrophils following AMI (**Supplementary Table 9)**.

### miRNA-126-mRNA targets are overrepresented in neutrophil transcriptomes following AMI

Human miRNA-126-mRNA targets were significantly overrepresented in human neutrophil transcriptomes at the time of injury (P<0.05) (**Supplementary Table 10**). There was no association between the differentially expressed genes in the mouse blood neutrophil sc-RNA-sequencing populations and miRNA-126-mRNA targets (**Supplementary Table 10**), but neutrophils within the myocardium displayed differential enrichment for miRNA-126-mRNA targets (**Supplementary Table 10**). There was significant enrichment for miRNA-126-mRNA targets from the human, mouse and the overlap lists (P<0.001), consistent with a role for miRNA-126 in regulating neutrophil gene expression following AMI.

To test the functional significance of these findings, we treated wild-type mice with an antagomiR for miRNA-126 (n = 5) or a scramble control (n = 7) prior to induction of experimental AMI to assess the role of miRNA-126 on infarct size (**Figure 4E**) and found a 12% reduction in infarct size in animals that received the miRNA-126 antagomir versus the scramble control (P<0.01) (**Figure 4F**).

### EC-EVs localise to the spleen

These accumulating data suggest that EC-EVs, enriched for miRNA-126 and VCAM-1 provide an ‘ischaemia signal’ to neutrophils in the spleen, resulting in mobilisation and transcriptional activation. Accordingly, we tested whether EC-EV localised to the spleen after intravenous injection and whether there were consequent alterations in neutrophil-associated chemokine gene and protein expression. Primary mouse and human endothelial cells release more EVs following inflammatory stimulation *(24)* and hypoxia *(40)*. In agreement with these data and our previous observations, mouse sEND.1 endothelial cells produced EV under basal conditions and released significantly more EVs after pro-inflammatory stimulation with TNF-α (P<0.001) (**Figure 5A/B)** *(24)*. We confirmed endothelial cell activation by TNF-α by probing for VCAM-1 protein and found significantly more protein (P<0.001) (**Figure 3C**). EC-EVs displayed typical EV morphology (**Figure 5D/E)**, EV-protein markers ALIX, TSG101 and CD9 **(Figure 5F)** and were positive for eNOS and VCAM-1. EV-depleted cell culture supernatants and cell culture media that had not been exposed to cells were negative for ALIX, TSG101, CD9, eNOS and VCAM-1, confirming robust EC-EV isolation. EC-EVs derived from pro-inflammatory stimulations showed significant enrichment for miRNA-126-3p (P<0.001) (**Figure 5G)** and miRNA-126-5p (P<0.01) (**Figure 5H)**, indicating similarities in the EC-EV response between human and mouse endothelial cells. We labelled the mouse EC-EV by transfection with non-mammalian miRNA-39, which belongs to *C. elegans* and allows quantitative tracing of EVs *in vivo* (**Figure 6A)**. This EC-EV labelling approach showed that EC-EVs accumulated rapidly in the spleen and were detectable 2 hours, 6 hours (P<0.001) and 24 hours (P<0.001) post-intravenous injection (**Figure 6A)**.

**Figure 5.**
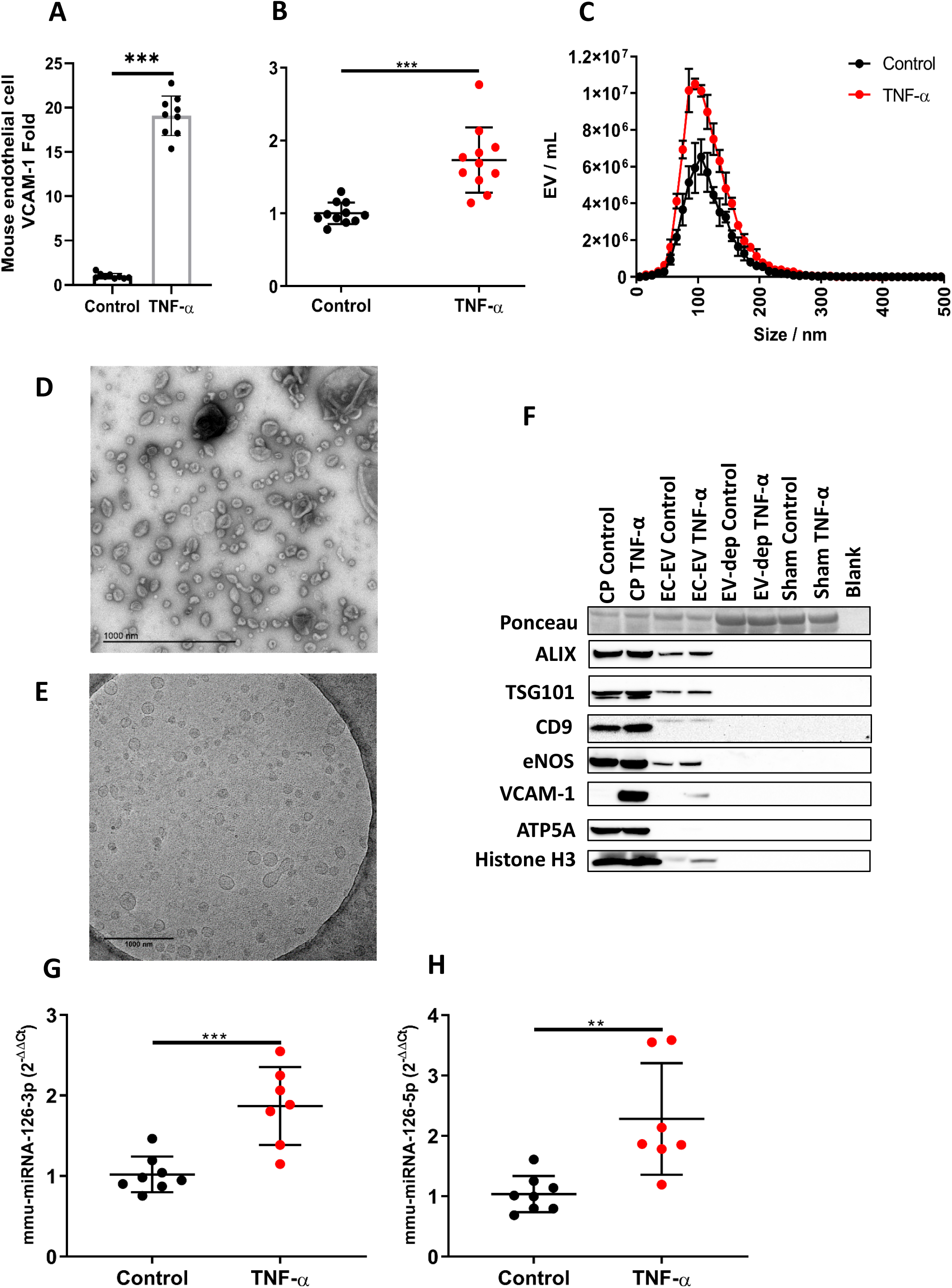
Mouse sEND.1 endothelial cells release more extracellular vesicles (EVs) after inflammatory stimulation. **A** mouse sEND.1 endothelial cells express more VCAM-1 following treatment with recombinant mouse tumour necrosis factor-α (TNF-α) (10 ng / mL) *(n* = 9 per group); **B** release more EVs *(n* = 11 per group). **C** Size and concentration profile of sEND.1 derived EVs under basal conditions *(n* = 3) and after inflammatory stimulation with recombinant mouse TNF-α *(n* = 4). **D** Transmission electron micrograph (TEM) of sEND.1-derived EVs (scale bar 1000 nm) and **E** cryo-TEM sEND.1-derived EVs (scale bar 1000 nm). **F** Ponceau stain and western blot of sEND.1 derived EV from basal and after inflammatory stimulation with TNF-α for ALIX, TSG101, CD9, eNOS, VCAM-1, ATP5A and Histone H3. sEND1 cell pellets (CP), EV-depleted cell culture supernatants (EV-dep) and cell culture media that was not exposed to cells (sham) were used as controls. EC-EV miRNA levels of **G** hsa-miRNA-126-3p and **H** hsa-miRNA-126-5p under basal conditions *(n* = 8) and after inflammatory stimulation with TNF-α *(n* = 7). An unpaired t test was used in **A, B, C, G** and **H** for statistical analysis. Error bars represent mean □± □SD \*\**p* □< □0.01, \*\*\**p* □< □0.001.

**Figure 6.**
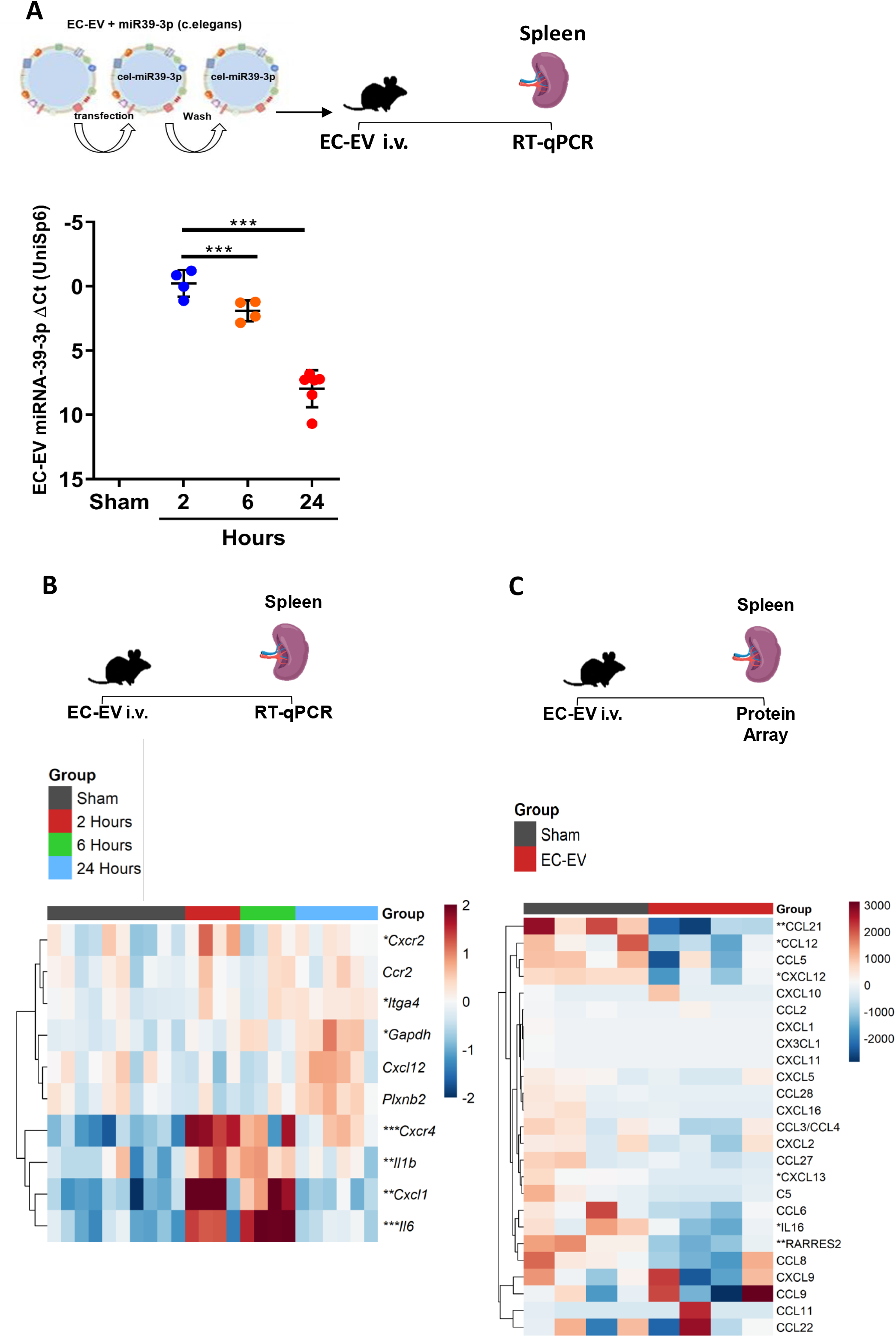
Endothelial cell derived extracellular vesicles (EC-EV) localise to the spleen in wild-type, mice and influence gene and protein expression. **A** RT-qPCR detection of EC-EV labelled with miRNA-39-3p in the spleen of mice following intravenous injection of 1 × 10 ^9^ EVs by tail vein at 2 *(n* = 4), 6 *(n* = 4) and 24 *(n* = 6) hours post-injection. Sham *(n* = 10) represents a media only preparation control with no EC-EVs. **B** Heat map showing gene expression in the spleen of mice following intravenous injection of 1 × 10 ^9^ EVs by tail vein at 2 *(n* = 4), 6 *(n* = 4) and 24 *(n* = 6) hours post injection. Sham *(n* = 10) represents a media only preparation control with no EC-EVs. Data shown as ΔΔCt values normalised to row mean ΔΔCt value for each gene. **C** Heat map showing protein expression in the spleen of mice following intravenous injection of 1 × 10 ^9^ EVs by tail vein at 2 hours *(n* = 4) post-injection. Sham *(n* = 4) represents a media only preparation control with no EC-EVs. Data shown are chemokine array dot blot density values normalised to mean row value for each protein. A one-way ANOVA with post-hoc Bonferroni correction was used in **A, B** and an unpaired t test was used in **C**. Error bars represent mean □± SD □ \*\**p* □< □0.05, \*\**p* □< □0.01, \*\*\**p* □< □0.001.

### EC-EVs alter chemokine and protein expression in the spleen

Informed by the earlier *in silico* studies suggesting regulation of neutrophil activation and motility by miRNA-126, we hypothesised that EC-EV localisation in the spleen would alter gene expression within spleen tissue, with a focus on CXC chemokine and cytokine activity.

EC-EVs induced significantly altered mRNA in expression for *Cxcr2, Itag4, Gapdh* (all, P<0.05), *Il-1*β, *Cxcl1* (both, P<0.01), *Cxcr4 and Il-6* (both, P<0.001) post-EC-EV injection **(Figure 6B)**.

We further determined whether delivery of EC-EVs to the spleen altered chemokine protein levels, including for the retention chemokine CXCL12 / SDF-1. In the same mice we undertook a quantitative protein-detection array for 25 different proteins that influence neutrophil function, including CXCL12 / SDF-1, CCL2 *(41)* and CCL3 *(42)*, which are known miRNA-126-mRNA targets and CCL27/ CCL28, which are predicted miRNA-126-mRNA targets. There were significant reductions in 6CKine/CCL21 (P<0.01), BLC/CXCL13 (P<0.05), chemerin/retinoic acid receptor responder protein 2 (RARRES2) (P<0.01), IL-16 (P<0.05), MCP-5/CCL12 (P<0.05) and CXCL12 / SDF-1. (P<0.05) **(Figure 6C)**. These findings are consistent with a role for EC-EV-miRNA-126 in silencing genes involved in cell retention.

### Endothelial cell derived EVs mobilise neutrophils from the spleen

Given the effects of the EC-EVs derived from TNF-α activated cells on gene expression and silencing of retention chemokines, we injected EC-EVs, derived from TNF-α activated endothelial cells, intravenously into healthy wild-type mice vs sham injection **(Figure 7A)**. Flow cytometry (Live, CD45^+^, CD11b^+^, Ly6G^+^) showed that EC-EVs significantly increased the number of circulating peripheral blood neutrophils **(Figure 7B)**, and simultaneously lowered splenic-neutrophil numbers in the same mice **(Figure 7B)**, confirming splenic neutrophil mobilisation induced by EC-EV. Consistent with our observations in AMI we found that EC-EVs mediated greater neutrophil mobilisation from the spleen (P<0.001) than from the bone marrow **(Figure 7C)**. As in the context of AMI, there was no alteration in CD62L/L-selectin expression in blood neutrophils **(Figure 7D)**.

**Figure 7.**
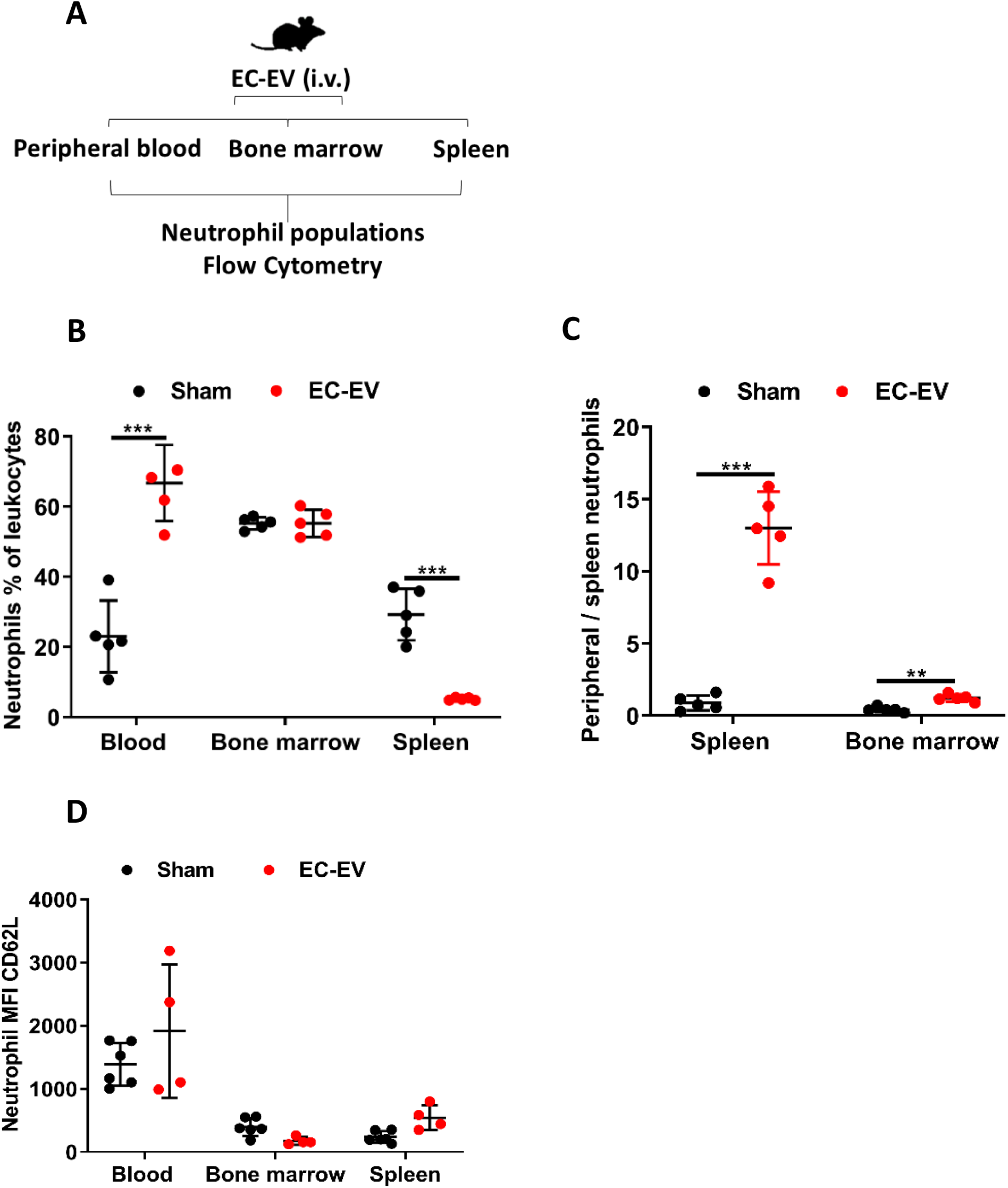
Endothelial cell derived extracellular vesicles (EC-EV) (1 × 10 ^9^ EVs / mL) mobilise splenic-neutrophils in healthy mice. **A** Percentage of neutrophils as a proportion of the total leukocytes (live, CD45^+^, CD11b^+^, Ly6G^+^) in peripheral blood, bone marrow and spleen *(n* = 5 per group). **B** Splenic-neutrophil mobilisation ratio (peripheral blood neutrophils/ spleen neutrophils) shows net contributions of neutrophil reserves to mobilised peripheral blood neutrophils following intravenous injections of EC-EV (1 × 10 ^9^ EVs m mL) injections *(n* = 5 per group). **C** Mean fluorescent intensity (MFI) of CD62L/L-selectin on neutrophils in peripheral blood, spleen and bone marrow 2 hours after *(n* = 5 per group) intravenous injections of EC-EV (1 × 10 ^9^ EVs / mL). An unpaired t test was used in **A, B** and **C**. Error bars represent mean □± SD \*\*\**p* □< □0.001.

### EC-EV VCAM-1 mediates neutrophil mobilisation

Plasma EVs present VCAM-1 on their surface (**Figure 8A**) and the number of VCAM-1 positive EVs increases after AMI *(24)*. Similarly, endothelial cells in culture produce EVs enriched for VCAM-1 following pro-inflammatory stimulation (**Figure 5E**). Given its well-established role in mediating interactions between activated vascular endothelium and circulating leukocytes, we hypothesised that VCAM-1 on EC-EVs might perform the converse role by mediating the capture of circulating EC-EVs by static neutrophils in the spleen. We used CRISPR-Cas9 base editing of endothelial cells to produce VCAM-1 deficient EC-EVs by introducing a stop codon in the VCAM-1 sequence. To confirm CRISPR-Cas9 editing of VCAM-1 from endothelial cells we stimulated wild-type (WT) and VCAM-1 knock-out (KO) cells with TNF-α. WT mouse endothelial cells expressed more VCAM-1 following inflammatory stimulation (P<0.001) (**Figure 8B/C**), whereas VCAM-1 KO cells did not express VCAM-1, confirming successful CRISPR-Cas9 base editing in endothelial cells. VCAM-1 KO cells released EC-EV under basal conditions similar to WT cells (**Figure 8D/E)** but VCAM-1 KO endothelial cells did not release more EVs following inflammatory stimulation with TNF-α (**Figure 8D/E)**. WT and VCAM-1 KO EC-EV were positive for EV-markers TSG101 and CD9 but only WT inflammatory derived EC-EVs were positive for VCAM-1 (**Figure 8F**). EC-EVs derived from either TNF-α activated WT or TNF-α activated VCAM-1 KO cells were injected into healthy, wild-type mice at the same concentration (1 × 10^9^ / mL EC-EVs). Using the miRNA-39-3p labelling technique (described above) we found that VCAM-1 deficient EC-EVs and WT EC-EV accumulate in the spleen at similar levels (**Figure 8G**), but VCAM-1 deficient EC-EVs did not induce alteration in gene expression that were comparable to WT EC-EVs responses in the spleen for *Il-6* (P<0.001), *Il-1*β (P<0.05) and *Cxcl1* (P<0.05) (**Figure 8H)**, suggesting alter signalling. Furthermore, deletion of VCAM-1 in EC-EVs prevented mobilisation of splenic neutrophils to peripheral blood when compared to wild-type VCAM-1+ EC-EVs (**Figure 8I and 8J)**.

**Figure 8.**
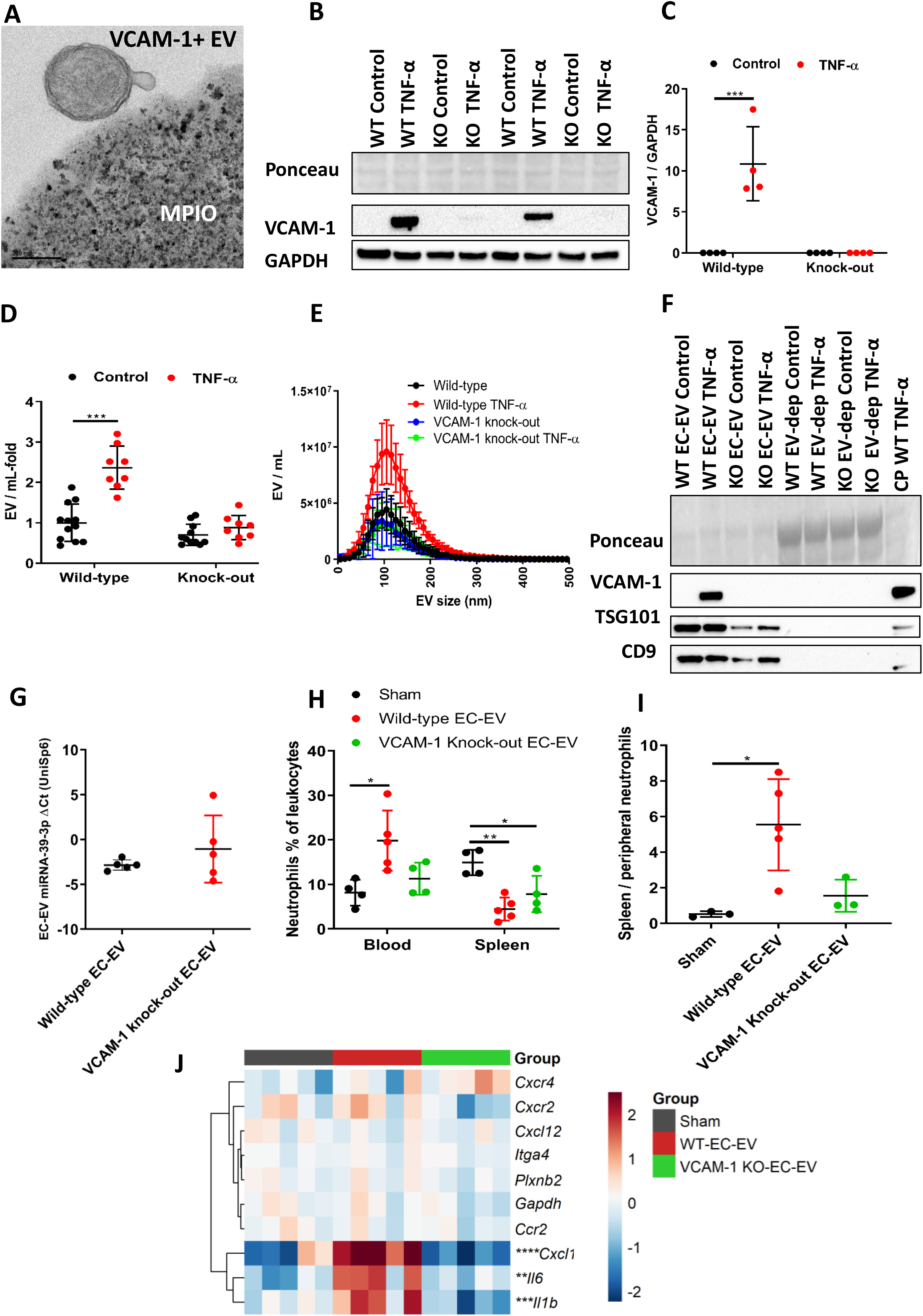
Extracellular vesicle (EV) vascular cell adhesion molecule-1 (VCAM-1) is necessary for endothelial cell (EC)-EV splenic-neutrophil mobilisation. **A** Transmission electron micrograph of a VCAM-1+ plasma EV bound to a magnetic bead of iron oxide (MPIO) conjugated with anti-human VCAM-1 antibodies, scale bar is 200nm. **B**/**C** Western blot of sEND.1 wild-type and CRISPR-cas9 base-edited VCAM-1 knock outs cell pellets under basal conditions (wild-type *n* = 4 and VCAM-1 knock out *n* = 4 per group) and after inflammatory stimulation with recombinant mouse tumour necrosis (TNF-α). **D** The number of mouse sEND.1 endothelial cell derived extracellular vesicles (EC-EVs) from wild-type and CRISPR-cas9 base-edited VCAM-1 knock outs under basal conditions (wild-type *n* = 12 and VCAM-1 knock out *n* = 11 per group) and after inflammatory stimulation with recombinant mouse tumour necrosis factor-α (TNF-α) *(n* = 8 per group). **E** Size and concentration profile of mouse sEND.1 EC-EVs from wild-type and CRISPR/Cas9 base-edited VCAM-1 knock outs under basal conditions (wild-type *n* = 12 and VCAM-1 knock out *n* = 11 per group) and after inflammatory stimulation with recombinant mouse tumour necrosis factor-α (TNF-α) *(n* = 8 per group). **F** Ponceau stain and western blot of wild-type and CRISPR-case9 base-edited VCAM-1 knock out sEND.1 derived EVs from basal and after inflammatory stimulation with recombinant mouse TNF-α for TSG101, CD9 and VCAM-1. Inflammatory stimulated s.END1 cell pellets and EV-depleted cell culture supernatants were used as controls. **G** RT-qPCR detection of wild-type sEND.1 and CRISPR-cas9 base-edited VCAM-1 knock outs EC-EV labelled with miRNA-39-3p in the spleen of mice following intravenous injection of 1 × 10 ^9^ EVs by tail vein at 2 hours post-injection *(n* = 5 per group). **H** Heat map showing mRNA expression in the spleen of mice following intravenous injection of wild-type or CRISPR-cas9 base-edited VCAM-1 knock out EC-EVs 1 × 10^9^ EVs by tail vein at 2 hours post-injection. Sham represents a media only preparation control with no EC-EVs *(n* = 5 per group). Data shown as ΔΔCt values normalised to row mean ΔΔCt value for each gene. **I/J** Percentage of neutrophils as a proportion of the total leukocytes (live, CD45^+^, CD11b^+^, Ly6G^+^) in peripheral blood and spleen (sham and VCAM-1 knock out EC-EV *n* = 4 and wild-type EC-EV *n* = 5 per group). **I** Splenic-neutrophil mobilisation ratio (peripheral blood neutrophils/ spleen neutrophils) shows net contributions of splenic reserves to mobilised peripheral blood neutrophils following intravenous injections of wild-type or CRISPR-cas9 base-edited VCAM-1 knock out EC-EVs 1 × 10^9^ EVs by tail vein at 2 hours post injection. Sham represents a media only preparation control with no EC-EVs (sham and VCAM-1 knock out EC-EV *n* = 3 and wild-type EC-EV *n* = 5 per group). One-way (**H, I** and **J**) and two-way (**B, C, D** and **E**) ANOVA with post-hoc Bonferroni correction was used for statistical analysis. An unpaired t test was used in **F**. Error bars represent mean □± □SD \**p* □< □0.05, \*\**p* □< □0.01, \*\*\**p* □< □0.001, \*\*\*\**p* □< □0.0001.

## Discussion

Mobilisation of neutrophils occurs rapidly after AMI in mice and humans *(1, 3, 4)* and their number in peripheral blood correlates with the extent of myocardial injury *(1-3)*. The bone marrow has been regarded as the principal source for neutrophils that are mobilised to peripheral blood after injury, because (i) it is the primary site for granulopoiesis *(8-10)*; (ii) it contains ample reserves of mature cells and; (iii) releases neutrophils in response to injection of exogenous chemokines *(12-16)*. However, the divergent timings of neutrophil mobilisation (rapid) *(1, 20)* and chemokine elevation (delayed) *in vivo (17, 18, 43)* suggest that additional processes may be involved.

Here, we have identified a previously unknown mechanism by which ischemic injury to the myocardium signals to mobilization of neutrophils from a splenic reserve. We show that: (i) EC-EV generated under conditions of inflammation are enriched for VCAM-1, miRNA-126-3p and miRNA-126-5p; (ii) EC-EVs are delivered to the spleen, where they alter gene and protein expression; and (iii) induce the mobilisation of splenic neutrophils to peripheral blood. Notably, (iv) these EC-EV effects are dependent on VCAM-1. Furthermore, (v) we show that neutrophil transcriptomes are differentially regulated following AMI, prior to entry into the myocardium. (vi) Targets of miRNA-126 are significantly altered in neutrophil transcriptomes post-AMI and (vii) administration of miRNA-126 antagomir significantly reduces infarct size *in vivo*.

Mature neutrophils are held in large numbers in the haemopoietic cords through interactions with the neutrophil receptors CXCR4 and CXCL12 *(11, 44, 45)*. Loss of CXCL12 induces an increase in peripheral blood neutrophils. Injection of chemotactic factors *(13)*, CXCL chemokines *(12, 14)* and G-CSF *(15, 16)* can drive the rapid mobilisation of neutrophils across the sinusoidal endothelium through alterations in CXCR4-CXCL12.

Scrutiny of the relative timings of cytokine elevation after ischaemic injury in relation to neutrophil mobilisation does not support their role in this early mobilisation, since both the onset and peaks in neutrophil mobilisation occur prior to those for cytokine elevation *(17, 18, 43)*. IL-8 injection mobilised neutrophils from the bone marrow *(13)*, but after reperfusion in AMI, even in blood from the coronary sinus (undiluted myocardial effluent), the elevation is modest (0.1-fold) *(18, 19)*. Furthermore, we calculate that the absolute concentration based on these physiological measurements is approximately 2-3 orders of magnitude less than the concentration used to elicit neutrophil mobilisation in mice *(14)*. Finally, neutrophils are the first cells to arrive in the acutely injured tissue. Neutrophil depletion dampens plasma chemokines levels following AMI *(7)* and in a mouse air pouch model *(46)*. It is not clear which other cells in the profoundly ischaemic myocardium could be capable of the rapid synthesis of chemokines, that would be of sufficient magnitude to mediate neutrophil mobilisation from a remote site such as the bone marrow. The sympathetic nervous system is also activated following AMI and mobilises hematopoietic stem cells (HSCs) from the bone marrow *(47)*. However, HSCs are immature cells unlike terminally differentiated neutrophils and blood numbers of HSCs do not peak until >6 hours post-AMI, subsequent to the increase in peripheral blood neutrophils.

By contrast, numerous studies have shown that hypoxia promotes the rapid (<30 minutes) release of EVs by endothelial cells *(40)*. We show that activated endothelial cells in culture liberate large amounts of EV that contain VCAM-1 in their membranes. Using CRISPR/Cas9 genome base editing of cultured endothelial cells, we generatedVCAM-1-deficient EV and showed that while VCAM-1 was not essential for splenic localisation, its absence removed the ability of EV to provoke the rapid mobilisation of neutrophils. Importantly, EVs are taken up rapidly and selectively by the spleen where they become locally concentrated *(24, 48)*, unlike chemokines, which have a systemic effect.

Following injury to the myocardium there is a marked increase in VCAM-1-bearing-EVs. VCAM-1 is a glycoprotein, which is expressed on activated endothelium and has a well-established role in the recruitment of circulating leukocytes by binding integrins *(31, 49, 50)*, including CD49d *(51)* and CD11b/CD18 *(52)*. Therefore, our findings suggest an efficient signalling system, in which neutrophils are activated and mobilised by engaging VCAM-1-bearing EVs that are taken up in the spleen, having been released remotely from activated endothelium. A subsequent interaction between neutrophils in circulation and static VCAM-1 on activated endothelial cells mediates their recruitment to the original site of injury.

The recruitment of neutrophils to the injured myocardium is an essential step in tissue response to injury and repair *(1, 7, 20)* and thus modulating the neutrophil response raises possibilities for immuno-modulatory interventions in selected inflammatory pathologies, including AMI. Peripheral blood neutrophils are elevated at time of presentation with AMI in patients and rapidly increase in peripheral blood following AMI in mice. Here, we show that splenic-neutrophils are rapidly mobilised to peripheral blood by EC-EV bearing VCAM-1. These findings complement the current paradigm in which neutrophils are liberated from bone marrow reserves through elevations in blood chemokines. We demonstrate a novel and efficient signalling mechanism between the injured heart microvasculature and the spleen.

Endothelial cells are ideally placed for the rapid release of EVs to peripheral blood during ischemia. EV clearance is rapid and predominately to the spleen, which contains neutrophils in the sup-capsular red pulp. Precisely how neutrophils are retained in the spleen is not known, but our findings suggest that local chemokine signalling may be important, as delivery of endothelial cell derived EVs rich in miRNA-126 downregulates retention chemokines, including the miRNA-126 target CXCL12, and induces expression of neutrophil mobilisation signals, namely CXCL1. Importantly, we show that these processes are dependent on EV-VCAM-1, an integrin with a well-documented role in immune cell recruitment. Furthermore, we make a new observation that peripheral blood neutrophils are transcriptionally activated prior to recruitment to the injured myocardium, with a bias towards miRNA-126-mRNA targets. Our bioinformatics analysis revealed SRP-dependent co-translational protein targeting to membrane as the most significantly enriched pathway that was conserved between the human and the mouse blood neutrophils following AMI. SRP is necessary for transferring newly synthesised nascent proteins from the ribosome, which are destined for cellular excretion. SRP monoallelic mutations induce a congenital blood neutropenia, possibly due to an increase in susceptibility of neutrophils to apoptosis and the unfolded protein response *(53, 54)*. A significant enrichment for the universally conserved SRP supports our conclusion that neutrophils are transcriptionally active prior to recruitment to the injured heart. Targeting SRP may open novel opportunities to target neutrophil transcriptomes prior to tissue recruitment to modulate their survival, function and phenotype, thereby protecting injured tissues from pro-inflammatory neutrophil mediated damage.

In conclusion, we demonstrate that the injured myocardium can rapidly mobilise splenic neutrophils through generation and release of EC-EVs that bear VCAM-1. These findings provide novel insights into how neutrophils are mobilised to peripheral blood following ischaemic injury, without the need for immediate generation and release of chemokines. EVs are decorated in surface proteins and integrins, which allows them to interact with cells and home to specific sites *(55, 56)*. A functionally efficient reciprocity may operate, in which VCAM-1, on endothelial cell-derived EVs is required for the mobilisation of splenic neutrophils, complementing the known role of static VCAM-1 in the recruitment of circulating neutrophils to activated endothelium. Neutrophils are the first cells recruited to the ischemic myocardium and are a major source of chemokines *(7)*. Thus, the well-established mobilisation of neutrophils from bone marrow reserves in response to chemokines *(12-16)* represent a secondary response, which is consistent with the time course of earlier reports and with our observations that there is no rapid bone marrow mobilisation in the early phase.

We have shown proof of concept that genetic manipulation can alter EV properties in functionally important ways. Immunomodulation of the neutrophil and monocyte response to AMI using EV vectors may provide therapeutic opportunities in AMI.

## Materials and Methods

### Acute myocardial infarction patients

All clinical investigations were conducted in accordance with the Declaration of Helsinki. The Oxfordshire Research Ethics Committee (references 08/H0603/41 and 11/SC/0397) approved human clinical cohort protocols. All patients provided informed written consent for inclusion in the study.

### LAD ligation model

All animal procedures were approved by an ethical review committee at the University of Oxford or NYU Lagone Health. UK experimental interventions were carried out by UK Home Office personal licence holders under the authority of a Home Office project licence.

Further experimental details are provided in the supplementary methods.

### Data Availability

RNA Sequencing data are deposited at Gene Expression Omnibus. *Pending* accession number.

### Statistical analysis

All values are group mean ± standard deviation (s.d.). Paired and unpaired two-tailed student’s t test was used to compare 2 groups, a one-way or two-way analysis of variance (ANOVA) or mixed model effects with post-hoc Bonferroni correction was used to compare multiple group (>2) means with one, two or more independent variables. *p* values <0.05 were considered significant. Hierarchical clustering analysis and generation of heatmap plots was performed using the pheatmap R package v1.0.12.

## Data Availability

Data Availability
RNA Sequencing data are deposited at Gene Expression Omnibus. Pending accession number.

## Statement of Contribution

NA isolated, characterized, and utilized EVs in the described experiments and performed western blots, RT-qPCR, chemokine arrays, analysis of infarcted hearts and assisted with *in silico* bioinformatics. ATB, CL, EL, LE, GM, EM, CvS assisted with experimentation. EEC, GJK and CvS undertook antagomiR experiments. ATB performed bioinformatics analysis and generated transcriptome analysis graphs and heat maps. ALC and DP prepared and analysed flow cytometry preparations. AC, RD and EJ imaged EVs by TEM /Cryo-TEM. MGH performed some AMI surgeries. TK and CB performed RNA sequencing. JR performed CRISPR-cas9 base editing of endothelial cells. KJM, PR, KM and DCA led animal investigations. RC and KMC led human AMI investigations. NA and RC conceived the study. All authors participated in study design, coordination and helped to draft the manuscript. All authors have seen the final version of the manuscript and approve of its submission.

The authors thank the staff at the Oxford Heart Centre for the clinical care of patients recruited in the coordination of the OxAMI study. Phil Townsend and Steve Woodhouse are gratefully acknowledged for general laboratory management. The authors thank biomedical services staff for their expert care of mice used in this study. The OxAMI study is supported by the British Heart Foundation (BHF; grant CH/16/1/32013 to KMC), the BHF Centre of Research Excellence, Oxford (RG/13/1/30181), the National Institute for Health Research Biomedical Research Centre, Oxford and the and the Biomedical Sequencing Facility (BSF) at CeMM for assistance with next-generation sequencing. See Supplemental Acknowledgments for OxAMI details.

## Figures and Figure Legends

**Supplementary Table 1** miRNA-126-mRNA targets in the human, mouse and the overlap between both species.

**Supplementary Table 2** miRNA-126-mRNA target pathways using gene ontology (GO) in the human, mouse and the overlap between both species.

**Supplementary Table 3** P-values are shown from a Fisher’s exact test used to detect enrichment of curated miRNA-126-mRNA targets versus gene sets for neutrophil GO terms.

**Supplementary Table 4** GO biological process and Reactome pathway analysis of differentially enriched genes in human neutrophils from STEMI patients at the time of injury.

**Supplementary Table 5** GO (biological process, molecular function and cellular component) and Reactome pathway analysis in single cell RNA-sequencing of differentially enriched genes in neutrophil blood population 1 follow AMI.

**Supplementary Table 6** GO (biological process, molecular function and cellular component) and Reactome pathway analysis in single cell RNA-sequencing of differentially enriched genes in neutrophil blood population 2 follow AMI.

**Supplementary Table 7** GO (biological process, molecular function and cellular component) and Reactome pathway analysis in single cell RNA-sequencing of differentially enriched genes in neutrophil blood population 3 follow AMI.

**Supplementary Table 8** P-values are shown from a Fisher’s exact test used to detect enrichment of differentially enriched genes in human peripheral blood neutrophil versus single cell RNA-sequencing of blood neutrophils from the mouse following AMI.

**Supplementary Table 9** P-values and Jaccard index are shown from a Fisher’s exact test used to detect enrichment of differentially enriched gene target pathways using GO (biological process, molecular function and cellular component) and Reactome pathway analysis from human peripheral blood neutrophil versus single cell RNA-sequencing of blood neutrophils from the mouse following AMI.

**Supplementary Table 10** miRNA-126-mRNA targets from the human, mouse and the overlap (present in human and mouse) were compared with differentially expressed (DE) genes and the not-DE (NDE) genes in the neutrophil transcriptomes at time of presentation with AMI.

miRNA-126-mRNA targets from the human, mouse and the overlap (present in human and mouse) were compared with differentially expressed (DE) genes and the not-DE (NDE) genes in single cell RNA-sequencing of differentially enriched genes in neutrophil blood and heart populations following AMI.

**Supplementary OxAMI**. Author acknowledgments for the Oxford Acute Myocardial Infarction Study.

**Supplementary Methods**. Details of experimental methods and reagents.

**Supplementary uncropped western blots**. Unedited images of western blot membranes

## References

1. M. J. Schloss, M. Horckmans, K. Nitz, J. Duchene, M. Drechsler, K. Bidzhekov, C. Scheiermann, C. Weber, O. Soehnlein, S. Steffens, The time-of-day of myocardial infarction onset affects healing through oscillations in cardiac neutrophil recruitment. EMBO Mol Med 8, 937–948 (2016).

2. T. M. Guo, B. Cheng, L. Ke, S. M. Guan, B. L. Qi, W. Z. Li, B. Yang, Prognostic Value of Neutrophil to Lymphocyte Ratio for In-hospital Mortality in Elderly Patients with Acute Myocardial Infarction. Curr Med Sci 38, 354–359 (2018).

3. T. Kong, T. H. Kim, Y. S. Park, S. P. Chung, H. S. Lee, J. H. Hong, J. W. Lee, J. S. You, I. Park, Usefulness of the delta neutrophil index to predict 30-day mortality in patients with ST segment elevation myocardial infarction. Scientific reports 7, 15718 (2017).

4. G. Sreejit, A. Abdel-Latif, B. Athmanathan, R. Annabathula, A. Dhyani, S. K. Noothi, G. A. Quaife-Ryan, A. Al-Sharea, G. Pernes, D. Dragoljevic, H. Lal, K. Schroder, B. Y. Hanaoka, C. Raman, M. B. Grant, J. E. Hudson, S. S. Smyth, E. R. Porrello, A. J. Murphy, P. R. Nagareddy, Neutrophil-Derived S100A8/A9 Amplify Granulopoiesis After Myocardial Infarction. Circulation 141, 1080–1094 (2020).

5. S. Chia, J. T. Nagurney, D. F. Brown, O. C. Raffel, F. Bamberg, F. Senatore, F. J. Wackers, I. K. Jang, Association of leukocyte and neutrophil counts with infarct size, left ventricular function and outcomes after percutaneous coronary intervention for ST-elevation myocardial infarction. The American journal of cardiology 103, 333–337 (2009).

6. H. Lorchner, J. Poling, P. Gajawada, Y. Hou, V. Polyakova, S. Kostin, J. M. Adrian-Segarra, T. Boettger, A. Wietelmann, H. Warnecke, M. Richter, T. Kubin, T. Braun, Myocardial healing requires Reg3beta-dependent accumulation of macrophages in the ischemic heart. Nature medicine 21, 353–362 (2015).

7. M. Horckmans, L. Ring, J. Duchene, D. Santovito, M. J. Schloss, M. Drechsler, C. Weber, O. Soehnlein, S. Steffens, Neutrophils orchestrate post-myocardial infarction healing by polarizing macrophages towards a reparative phenotype. European Heart Journal 38, 187–197 (2016).

8. M. Evrard, I. W. H. Kwok, S. Z. Chong, K. W. W. Teng, E. Becht, J. Chen, J. L. Sieow, H. L. Penny, G. C. Ching, S. Devi, J. M. Adrover, J. L. Y. Li, K. H. Liong, L. Tan, Z. Poon, S. Foo, J. W. Chua, I. H. Su, K. Balabanian, F. Bachelerie, S. K. Biswas, A. Larbi, W. Y. K. Hwang, V. Madan, H. P. Koeffler, S. C. Wong, E. W. Newell, A. Hidalgo, F. Ginhoux, L. G. Ng, Developmental Analysis of Bone Marrow Neutrophils Reveals Populations Specialized in Expansion, Trafficking, and Effector Functions. Immunity 48, 364–379 e368 (2018).

9. Y. P. Zhu, L. Padgett, H. Q. Dinh, P. Marcovecchio, A. Blatchley, R. Wu, E. Ehinger, C. Kim, Z. Mikulski, G. Seumois, A. Madrigal, P. Vijayanand, C. C. Hedrick,Identification of an Early Unipotent Neutrophil Progenitor with Protumoral Activity in Mouse and Human Bone Marrow. Cell Rep 24, 2329–2341 e2328 (2018).

10. M. Horckmans, M. Bianchini, D. Santovito, R. T. A. Megens, J. Y. Springael, I. Negri, M. Vacca, M. Di Eusanio, A. Moschetta, C. Weber, J. Duchene, S. Steffens, Pericardial Adipose Tissue Regulates Granulopoiesis, Fibrosis, and Cardiac Function After Myocardial Infarction. Circulation 137, 948–960 (2018).

11. R. C. Furze, S. M. Rankin, Neutrophil mobilization and clearance in the bone marrow. Immunology 125, 281–288 (2008).

12. C. Martin, P. C. E. Burdon, G. Bridger, J. C. Gutierrez-Ramos, T. J. Williams, S. M. Rankin, Chemokines acting via CXCR2 and CXCR4 control the release of neutrophils from the bone marrow and their return following senescence. Immunity 19, 583–593 (2003).

13. T. Terashima, D. English, J. C. Hogg, S. F. van Eeden, Release of polymorphonuclear leukocytes from the bone marrow by interleukin-8. Blood 92, 1062–1069 (1998).

14. M. A. Jagels, T. E. Hugli, Neutrophil chemotactic factors promote leukocytosis. A common mechanism for cellular recruitment from bone marrow. Journal of immunology 148, 1119–1128 (1992).

15. C. L. Semerad, F. L. Liu, A. D. Gregory, K. Stumpf, D. C. Link, G-CSF is an essential regulator of neutrophil trafficking from the bone marrow to the blood. Immunity 17, 413–423 (2002).

16. A. Köhler, K. De Filippo, M. Hasenberg, C. van den Brandt, E. Nye, M. P. Hosking, T. E. Lane, L. Männ, R. M. Ransohoff, A. E. Hauser, O. Winter, B. Schraven, H. Geiger, N. Hogg, M. Gunzer, G-CSF-mediated thrombopoietin release triggers neutrophil motility and mobilization from bone marrow via induction of Cxcr2 ligands. Blood 117, 4349–4357 (2011).

17. A. Deten, H. C. Volz, W. Briest, H. G. Zimmer, Cardiac cytokine expression is upregulated in the acute phase after myocardial infarction. Experimental studies in rats. Cardiovascular research 55, 329–340 (2002).

18. F. J. Neumann, I. Ott, M. Gawaz, G. Richardt, H. Holzapfel, M. Jochum, A. Schomig, Cardiac release of cytokines and inflammatory responses in acute myocardial infarction. Circulation 92, 748–755 (1995).

19. N. Marx, F.-J. Neumann, I. Ott, M. Gawaz, W. Koch, T. Pinkau, A. Schömig, Induction of Cytokine Expression in Leukocytes in Acute Myocardial Infarction. Journal of the american college of cardiology 30, 165–170 (1997).

20. M. Horckmans, L. Ring, J. Duchene, D. Santovito, M. J. Schloss, M. Drechsler, C. Weber, O. Soehnlein, S. Steffens, Neutrophils orchestrate post-myocardial infarction healing by polarizing macrophages towards a reparative phenotype. Eur Heart J 38, 187–197 (2017).

21. I. Puga, M. Cols, C. M. Barra, B. He, L. Cassis, M. Gentile, L. Comerma, A. Chorny, M. Shan, W. Xu, G. Magri, D. M. Knowles, W. Tam, A. Chiu, J. B. Bussel, S. Serrano, J. A. Lorente, B. Bellosillo, J. Lloreta, N. Juanpere, F. Alameda, T. Baro, C. D. de Heredia, N. Toran, A. Catala, M. Torrebadell, C. Fortuny, V. Cusi, C. Carreras, G. A. Diaz, J. M. Blander, C. M. Farber, G. Silvestri, C. Cunningham-Rundles, M. Calvillo, C. Dufour, L. D. Notarangelo, V. Lougaris, A. Plebani, J. L. Casanova, S. C. Ganal, A. Diefenbach, J. I. Arostegui, M. Juan, J. Yague, N. Mahlaoui, J. Donadieu, K. Chen, A. Cerutti, B cell-helper neutrophils stimulate the diversification and production of immunoglobulin in the marginal zone of the spleen. Nat Immunol 13, 170–180 (2011).

22. J. F. Deniset, B. G. Surewaard, W. Y. Lee, P. Kubes, Splenic Ly6G(high) mature and Ly6G(int) immature neutrophils contribute to eradication of S. pneumoniae. J Exp Med 214, 1333–1350 (2017).

23. F. K. Swirski, M. Nahrendorf, M. Etzrodt, M. Wildgruber, V. Cortez-Retamozo, P. Panizzi, J. L. Figueiredo, R. H. Kohler, A. Chudnovskiy, P. Waterman, E. Aikawa, T. R. Mempel, P. Libby, R. Weissleder, M. J. Pittet, Identification of splenic reservoir monocytes and their deployment to inflammatory sites. Science 325, 612–616 (2009).

24. N. Akbar, J. E. Digby, T. J. Cahill, A. N. Tavare, A. L. Corbin, S. Saluja, S. Dawkins, L. Edgar, N. Rawlings, K. Ziberna, E. McNeill, S. Oxford Acute Myocardial Infarction, E. Johnson, A. A. Aljabali, R. A. Dragovic, M. Rohling, T. G. Belgard, I. A. Udalova, D. R. Greaves, K. M. Channon, P. R. Riley, D. C. Anthony, R. P. Choudhury, Endothelium-derived extracellular vesicles promote splenic monocyte mobilization in myocardial infarction. JCI Insight 2, 7;2(17):e93344 (2017).

25. C. Thery, K. W. Witwer, E. Aikawa, M. J. Alcaraz, J. D. Anderson, R. Andriantsitohaina, A. Antoniou, T. Arab, F. Archer, G. K. Atkin-Smith, D. C. Ayre, J. M. Bach, D. Bachurski, H. Baharvand, L. Balaj, S. Baldacchino, N. N. Bauer, A. A. Baxter, M. Bebawy, C. Beckham, A. Bedina Zavec, A. Benmoussa, A. C. Berardi, P. Bergese, E. Bielska, C. Blenkiron, S. Bobis-Wozowicz, E. Boilard, W. Boireau, A. Bongiovanni, F. E. Borras, S. Bosch, C. M. Boulanger, X. Breakefield, A. M. Breglio, M. A. Brennan, D. R. Brigstock, A. Brisson, M. L. Broekman, J. F. Bromberg, P. Bryl-Gorecka, S. Buch, A. H. Buck, D. Burger, S. Busatto, D. Buschmann, B. Bussolati, E. I. Buzas, J. B. Byrd, G. Camussi, D. R. Carter, S. Caruso, L. W. Chamley, Y. T. Chang, C. Chen, S. Chen, L. Cheng, A. R. Chin, A. Clayton, S. P. Clerici, A. Cocks, E. Cocucci, R. J. Coffey, A. Cordeiro-da-Silva, Y. Couch, F. A. Coumans, B. Coyle, R. Crescitelli, M. F. Criado, C. D’Souza-Schorey, S. Das, A. Datta Chaudhuri, P. de Candia, E. F. De Santana, O. De Wever, H. A. Del Portillo, T. Demaret, S. Deville, A. Devitt, B. Dhondt, D. Di Vizio, L. C. Dieterich, V. Dolo, A. P. Dominguez Rubio, M. Dominici, M. R. Dourado, T. A. Driedonks, F. V. Duarte, H. M. Duncan, R. M. Eichenberger, K. Ekstrom, S. El Andaloussi, C. Elie-Caille, U. Erdbrugger, J. M. Falcon-Perez, F. Fatima, J. E. Fish, M. Flores-Bellver, A. Forsonits, A. Frelet-Barrand, F. Fricke, G. Fuhrmann, S. Gabrielsson, A. Gamez-Valero, C. Gardiner, K. Gartner, R. Gaudin, Y. S. Gho, B. Giebel, C. Gilbert, M. Gimona, I. Giusti, D. C. Goberdhan, A. Gorgens, S. M. Gorski, D. W. Greening, J. C. Gross, A. Gualerzi, G. N. Gupta, D. Gustafson, A. Handberg, R. A. Haraszti, P. Harrison, H. Hegyesi, A. Hendrix, A. F. Hill, F. H. Hochberg, K. F. Hoffmann, B. Holder, H. Holthofer, B. Hosseinkhani, G. Hu, Y. Huang, V. Huber, S. Hunt, A. G. Ibrahim, T. Ikezu, J. M. Inal, M. Isin, A. Ivanova, H. K. Jackson, S. Jacobsen, S. M. Jay, M. Jayachandran, G. Jenster, L. Jiang, S. M. Johnson, J. C. Jones, A. Jong, T. Jovanovic-Talisman, S. Jung, R. Kalluri, S. I. Kano, S. Kaur, Y. Kawamura, E. T. Keller, D. Khamari, E. Khomyakova, A. Khvorova, P. Kierulf, K. P. Kim, T. Kislinger, M. Klingeborn, D. J. Klinke, 2nd, M. Kornek, M. M. Kosanovic, A. F. Kovacs, E. M. Kramer-Albers, S. Krasemann, M. Krause, I. V. Kurochkin, G. D. Kusuma, S. Kuypers, S. Laitinen, S. M. Langevin, L. R. Languino, J. Lannigan, C. Lasser, L. C. Laurent, G. Lavieu, E. Lazaro-Ibanez, S. Le Lay, M. S. Lee, Y. X. F. Lee, D. S. Lemos, M. Lenassi, A. Leszczynska, I. T. Li, K. Liao, S. F. Libregts, E. Ligeti, R. Lim, S. K. Lim, A. Line, K. Linnemannstons, A. Llorente, C. A. Lombard, M. J. Lorenowicz, A. M. Lorincz, J. Lotvall, J. Lovett, M. C. Lowry, X. Loyer, Q. Lu, B. Lukomska, T. R. Lunavat, S. L. Maas, H. Malhi, A. Marcilla, J. Mariani, J. Mariscal, E. S. Martens-Uzunova, L. Martin-Jaular, M. C. Martinez, V. R. Martins, M. Mathieu, S. Mathivanan, M. Maugeri, L. K. McGinnis, M. J. McVey, D. G. Meckes, Jr., K. L. Meehan, I. Mertens, V. R. Minciacchi, A. Moller, M. Moller Jorgensen, A. Morales-Kastresana, J. Morhayim, F. Mullier, M. Muraca, L. Musante, V. Mussack, D. C. Muth, K. H. Myburgh, T. Najrana, M. Nawaz, I. Nazarenko, P. Nejsum, C. Neri, T. Neri, R. Nieuwland, L. Nimrichter, J. P. Nolan, E. N. Nolte-‘t Hoen, N. Noren Hooten, L. O’Driscoll, T. O’Grady, A. O’Loghlen, T. Ochiya, M. Olivier, A. Ortiz, L. A. Ortiz, X. Osteikoetxea, O. Ostergaard, M. Ostrowski, J. Park, D. M. Pegtel, H. Peinado, F. Perut, M. W. Pfaffl, D. G. Phinney, B. C. Pieters, R. C. Pink, D. S. Pisetsky, E. Pogge von Strandmann, I. Polakovicova, I. K. Poon, B. H. Powell, I. Prada, L. Pulliam, P. Quesenberry, A. Radeghieri, R. L. Raffai, S. Raimondo, J. Rak, M. I. Ramirez, G. Raposo, M. S. Rayyan, N. Regev-Rudzki, F. L. Ricklefs, P. D. Robbins, D. D. Roberts, S. C. Rodrigues, E. Rohde, S. Rome, K. M. Rouschop, A. Rughetti, A. E. Russell, P. Saa, S. Sahoo, E. Salas-Huenuleo, C. Sanchez, J. A. Saugstad, M. J. Saul, R. M. Schiffelers, R. Schneider, T. H. Schoyen, A. Scott, E. Shahaj, S. Sharma, O. Shatnyeva, F. Shekari, G. V. Shelke, A. K. Shetty, K. Shiba, P. R. Siljander, A. M. Silva, A. Skowronek, O. L. Snyder, 2nd, R. P. Soares, B. W. Sodar, C. Soekmadji, J. Sotillo, P. D. Stahl, W. Stoorvogel, S. L. Stott, E. F. Strasser, S. Swift, H. Tahara, M. Tewari, K. Timms, S. Tiwari, R. Tixeira, M. Tkach, W. S. Toh, R. Tomasini, A. C. Torrecilhas, J. P. Tosar, V. Toxavidis, L. Urbanelli, P. Vader, B. W. van Balkom, S. G. van der Grein, J. Van Deun, M. J. van Herwijnen, K. Van Keuren-Jensen, G. van Niel, M. E. van Royen, A. J. van Wijnen, M. H. Vasconcelos, I. J. Vechetti, Jr., T. D. Veit, L. J. Vella, E. Velot, F. J. Verweij, B. Vestad, J. L. Vinas, T. Visnovitz, K. V. Vukman, J. Wahlgren, D. C. Watson, M. H. Wauben, A. Weaver, J. P. Webber, V. Weber, A. M. Wehman, D. J. Weiss, J. A. Welsh, S. Wendt, A. M. Wheelock, Z. Wiener, L. Witte, J. Wolfram, A. Xagorari, P. Xander, J. Xu, X. Yan, M. Yanez-Mo, H. Yin, Y. Yuana, V. Zappulli, J. Zarubova, V. Zekas, J. Y. Zhang, Z. Zhao, L. Zheng, A. R. Zheutlin, A. M. Zickler, P. Zimmermann, A. M. Zivkovic, D. Zocco, E. K. Zuba-Surma, Minimal information for studies of extracellular vesicles 2018 (MISEV2018): a position statement of the International Society for Extracellular Vesicles and update of the MISEV2014 guidelines. J Extracell Vesicles 7, 1535750 (2018).

26. N. Akbar, V. Azzimato, R. P. Choudhury, M. Aouadi, Extracellular vesicles in metabolic disease. Diabetologia 62, 2179–2187 (2019).

27. J. P. G. Sluijter, S. M. Davidson, C. M. Boulanger, E. I. Buzás, D. P. V. de Kleijn, F. B. Engel, Z. Giricz, D. J. Hausenloy, R. Kishore, S. Lecour, J. Leor, R. Madonna, C. Perrino, F. Prunier, S. Sahoo, R. M. Schiffelers, R. Schulz, L. W. Van Laake, K. Ytrehus, P. Ferdinandy, Extracellular vesicles in diagnostics and therapy of the ischaemic heart: Position Paper from the Working Group on Cellular Biology of the Heart of the European Society of Cardiology. Cardiovascular research 114, 19–34 (2018).

28. A. E. Russell, A. Sneider, K. W. Witwer, P. Bergese, S. N. Bhattacharyya, A. Cocks, E. Cocucci, U. Erdbrugger, J. M. Falcon-Perez, D. W. Freeman, T. M. Gallagher, S. Hu, Y. Huang, S. M. Jay, S. I. Kano, G. Lavieu, A. Leszczynska, A. M. Llorente, Q. Lu, V. Mahairaki, D. C. Muth, N. Noren Hooten, M. Ostrowski, I. Prada, S. Sahoo, T. H. Schoyen, L. Sheng, D. Tesch, G. Van Niel, R. E. Vandenbroucke, F. J. Verweij, A. V. Villar, M. Wauben, A. M. Wehman, H. Yin, D. R. F. Carter, P. Vader, Biological membranes in EV biogenesis, stability, uptake, and cargo transfer: an ISEV position paper arising from the ISEV membranes and EVs workshop. J Extracell Vesicles 8, 1684862 (2019).

29. X. Loyer, I. Zlatanova, C. Devue, M. Yin, K. Y. Howangyin, P. Klaihmon, C. L. Guerin, M. Kheloufi, J. Vilar, K. Zannis, B. K. Fleischmann, D. W. Hwang, J. Park, H. Lee, P. Menasche, J. S. Silvestre, C. M. Boulanger, Intra-Cardiac Release of Extracellular Vesicles Shapes Inflammation Following Myocardial Infarction. Circulation research 123, 100–106 (2018).

30. M. Cheng, J. Yang, X. Zhao, E. Zhang, Q. Zeng, Y. Yu, L. Yang, B. Wu, G. Yi, X. Mao, K. Huang, N. Dong, M. Xie, N. A. Limdi, S. D. Prabhu, J. Zhang, G. Qin, Circulating myocardial microRNAs from infarcted hearts are carried in exosomes and mobilise bone marrow progenitor cells. Nature communications 10, 959 (2019).

31. E. A. Ross, M. R. Douglas, S. H. Wong, E. J. Ross, S. J. Curnow, G. B. Nash, E. Rainger, D. Scheel-Toellner, J. M. Lord, M. Salmon, C. D. Buckley, Interaction between integrin alpha9beta1 and vascular cell adhesion molecule-1 (VCAM-1) inhibits neutrophil apoptosis. Blood 107, 1178–1183 (2006).

32. N. Ruparelia, J. E. Digby, A. Jefferson, D. J. Medway, S. Neubauer, C. A. Lygate, R. P. Choudhury, Myocardial infarction causes inflammation and leukocyte recruitment at remote sites in the myocardium and in the renal glomerulus. Inflamm Res 62, 515–525 (2013).

33. A. M. Akhtar, J. E. Schneider, S. J. Chapman, A. Jefferson, J. E. Digby, K. Mankia, Y. Chen, M. A. McAteer, K. J. Wood, R. P. Choudhury, In vivo quantification of VCAM-1 expression in renal ischemia reperfusion injury using non-invasive magnetic resonance molecular imaging. PloS one 5, e12800 (2010).

34. L. Osborn, C. Hession, R. Tizard, C. Vassallo, S. Luhowskyj, G. Chirosso, R. Lobb, Direct Expression Cloning of Vascular Cell-Adhesion Molecule-1, a Cytokine-Induced Endothelial Protein That Binds to Lymphocytes. Cell 59, 1203–1211 (1989).

35. V. Agarwal, G. W. Bell, J. W. Nam, D. P. Bartel, Predicting effective microRNA target sites in mammalian mRNAs. Elife 4, (2015).

36. C. Sticht, C. De La Torre, A. Parveen, N. Gretz, miRWalk: An online resource for prediction of microRNA binding sites. PloS one 13, e0206239 (2018).

37. W. Liu, X. Wang, Prediction of functional microRNA targets by integrative modeling of microRNA binding and target expression data. Genome Biol 20, 18 (2019).

38. B. Jassal, L. Matthews, G. Viteri, C. Gong, P. Lorente, A. Fabregat, K. Sidiropoulos, J. Cook, M. Gillespie, R. Haw, F. Loney, B. May, M. Milacic, K. Rothfels, C. Sevilla, V. Shamovsky, S. Shorser, T. Varusai, J. Weiser, G. Wu, L. Stein, H. Hermjakob, P. D’Eustachio, The reactome pathway knowledgebase. Nucleic acids research 48, D498–D503 (2019).

39. E. Vafadarnejad, G. Rizzo, L. Krampert, P. Arampatzi, A. P. Arias-Loza, Y. Nazzal, A. Rizakou, T. Knochenhauer, S. R. Bandi, V. A. Nugroho, D. J. J. Schulz, M. Roesch, P. Alayrac, J. Vilar, J. S. Silvestre, A. Zernecke, A. E. Saliba, C. Cochain, Dynamics of Cardiac Neutrophil Diversity in Murine Myocardial Infarction. Circulation research 127, e232–e249 (2020).

40. S. M. Davidson, J. A. Riquelme, Y. Zheng, J. M. Vicencio, S. Lavandero, D. M. Yellon, Endothelial cells release cardioprotective exosomes that may contribute to ischaemic preconditioning. Scientific reports 8, 15885 (2018).

41. E. Arner, N. Mejhert, A. Kulyte, P. J. Balwierz, M. Pachkov, M. Cormont, S. Lorente-Cebrian, A. Ehrlund, J. Laurencikiene, P. Heden, K. Dahlman-Wright, J. F. Tanti, Y. Hayashizaki, M. Ryden, I. Dahlman, E. van Nimwegen, C. O. Daub, P. Arner, Adipose tissue microRNAs as regulators of CCL2 production in human obesity. Diabetes 61, 1986–1993 (2012).

42. J. Agudo, A. Ruzo, N. Tung, H. Salmon, M. Leboeuf, D. Hashimoto, C. Becker, L. A. Garrett-Sinha, A. Baccarini, M. Merad, B. D. Brown, The miR-126-VEGFR2 axis controls the innate response to pathogen-associated nucleic acids. Nat Immunol 15, 54–62 (2014).

43. V. Rusinkevich, Y. Huang, Z.-y. Chen, W. Qiang, Y.-g. Wang, Y.-f. Shi, H.-t. Yang, Temporal dynamics of immune response following prolonged myocardial ischemia/reperfusion with and without cyclosporine A. Acta Pharmacologica Sinica 40, 1168–1183 (2019).

44. K. J. Eash, J. M. Means, D. W. White, D. C. Link, CXCR4 is a key regulator of neutrophil release from the bone marrow under basal and stress granulopoiesis conditions. Blood 113, 4711–4719 (2009).

45. B. T. Suratt, J. M. Petty, S. K. Young, K. C. Malcolm, J. G. Lieber, J. A. Nick, J. A. Gonzalo, P. M. Henson, G. S. Worthen, Role of the CXCR4/SDF-1 chemokine axis in circulating neutrophil homeostasis. Blood 104, 565–571 (2004).

46. E. Garcia-Ramallo, T. Marques, N. Prats, J. Beleta, S. L. Kunkel, N. Godessart, Resident cell chemokine expression serves as the major mechanism for leukocyte recruitment during local inflammation. J Immunol 169, 6467–6473 (2002).

47. P. Dutta, G. Courties, Y. Wei, F. Leuschner, R. Gorbatov, C. S. Robbins, Y. Iwamoto, B. Thompson, A. L. Carlson, T. Heidt, M. D. Majmudar, F. Lasitschka, M. Etzrodt, P. Waterman, M. T. Waring, A. T. Chicoine, A. M. van der Laan, H. W. Niessen, J. J. Piek, B. B. Rubin, J. Butany, J. R. Stone, H. A. Katus, S. A. Murphy, D. A. Morrow, M. S. Sabatine, C. Vinegoni, M. A. Moskowitz, M. J. Pittet, P. Libby, C. P. Lin, F. K. Swirski, R. Weissleder, M. Nahrendorf, Myocardial infarction accelerates atherosclerosis. Nature 487, 325–329 (2012).

48. S. C. Saunderson, A. C. Dunn, P. R. Crocker, A. D. McLellan, CD169 mediates the capture of exosomes in spleen and lymph node. Blood 123, 208–216 (2014).

49. J. V. Forrester, J. M. Lackie, Adhesion of neutrophil leucocytes under conditions of flow. Journal of cell science 70, 93–110 (1984).

50. Y. Taooka, J. Chen, T. Yednock, D. Sheppard, The integrin alpha9beta1 mediates adhesion to activated endothelial cells and transendothelial neutrophil migration through interaction with vascular cell adhesion molecule-1. The Journal of cell biology 145, 413–420 (1999).

51. T. B. Issekutz, M. Miyasaka, A. C. Issekutz, Rat blood neutrophils express very late antigen 4 and it mediates migration to arthritic joint and dermal inflammation. J Exp Med 183, 2175–2184 (1996).

52. A. C. Issekutz, L. Ayer, M. Miyasaka, T. B. Issekutz, Treatment of established adjuvant arthritis in rats with monoclonal antibody to CD18 and very late activation antigen-4 integrins suppresses neutrophil and T-lymphocyte migration to the joints and improves clinical disease. Immunology 88, 569–576 (1996).

53. C. Bellanné-Chantelot, B. Schmaltz-Panneau, C. Marty, O. Fenneteau, I. Callebaut, S. Clauin, A. Docet, G.-L. Damaj, T. Leblanc, I. Pellier, C. Stoven, S. Souquere, I. Antony-Debré, B. Beaupain, N. Aladjidi, V. Barlogis, F. Bauduer, P. Bensaid, O. Boespflug-Tanguy, C. Berger, Y. Bertrand, L. Carausu, C. Fieschi, C. Galambrun, A. Schmidt, H. Journel, F. Mazingue, B. Nelken, T. C. Quah, E. Oksenhendler, M. Ouachée, M. Pasquet, V. Saada, F. Suarez, G. Pierron, W. Vainchenker, I. Plo, J. Donadieu, Mutations in the SRP54 gene cause severe congenital neutropenia as well as Shwachman-Diamond-like syndrome. Blood 132, 1318–1331 (2018).

54. Y. Mizoguchi, S. Hesse, M. Linder, N. Zietara, M. Lyszkiewicz, Y. Liu, M. Tatematsu, P. Grabowski, K. Ahomaa, T. Jeske, S. Hollizeck, E. Rusha, M. K. Saito, M. Kobayashi, Z. Alizadeh, Z. Pourpak, S. Iurian, N. Rezaei, E. Unal, M. Drukker, B. Walzog, F. Hauck, J. Rappsilber, C. Klein, Defects in Signal Recognition Particle (SRP) Components Reveal an Essential and Non-Redundant Role for Granule Biogenesis and Differentiation of Neutrophil Granulocytes. Blood 134, 216–216 (2019).

55. A. Hoshino, B. Costa-Silva, T. L. Shen, G. Rodrigues, A. Hashimoto, M. Tesic Mark, H. Molina, S. Kohsaka, A. Di Giannatale, S. Ceder, S. Singh, C. Williams, N. Soplop, K. Uryu, L. Pharmer, T. King, L. Bojmar, A. E. Davies, Y. Ararso, T. Zhang, H. Zhang, J. Hernandez, J. M. Weiss, V. D. Dumont-Cole, K. Kramer, L. H. Wexler, A. Narendran, G. K. Schwartz, J. H. Healey, P. Sandstrom, K. J. Labori, E. H. Kure, P. M. Grandgenett, M. A. Hollingsworth, M. de Sousa, S. Kaur, M. Jain, K. Mallya, S. K. Batra, W. R. Jarnagin, M. S. Brady, O. Fodstad, V. Muller, K. Pantel, A. J. Minn, M. J. Bissell, B. A. Garcia, Y. Kang, V. K. Rajasekhar, C. M. Ghajar, I. Matei, H. Peinado, J. Bromberg, D. Lyden, Tumour exosome integrins determine organotropic metastasis. Nature 527, 329–335 (2015).

56. G. Rodrigues, A. Hoshino, C. M. Kenific, I. R. Matei, L. Steiner, D. Freitas, H. S. Kim, P. R. Oxley, I. Scandariato, I. Casanova-Salas, J. Dai, C. R. Badwe,B. Gril, M. Tesic Mark, B. D. Dill, H. Molina, H. Zhang, A. Benito-Martin, L. Bojmar, Y. Ararso, K. Offer, Q. LaPlant, W. Buehring, H. Wang, X. Jiang, T. M. Lu, Y. Liu, J. K. Sabari, S. J. Shin, N. Narula, P. S. Ginter, V. K. Rajasekhar, J. H. Healey, E. Meylan, B. Costa-Silva, S. E. Wang, S. Rafii, N. K. Altorki, C. M. Rudin, D. R. Jones, P. S. Steeg, H. Peinado, C. M. Ghajar, J. Bromberg, M. de Sousa, D. Pisapia, D. Lyden, Tumour exosomal CEMIP protein promotes cancer cell colonization in brain metastasis. Nat Cell Biol 21, 1403–1412 (2019).

