## Supplemental Methods for "Endothelial Cell-Derived Extracellular Vesicles Elicit Neutrophil Deployment from the Spleen Following Acute Myocardial Infarction"

Acute myocardial infarction (AMI) patients

A subset of patients that were previously recruited to the Oxford Heart Centre as part of the Oxford Acute Myocardial Infarction Study (OxAMI) for the analysis of plasma extracellular vesicles (EVs) and AMI were used in this study (*1*). Patients were selected by identifying neutrophil counts (10^9^ cells / L) in their clinical records (N = 15). Associations between plasma EV numbers, % of myocardial injury/oedema, scar formation by (LGE) were determined by CMR imaging and plasma neutrophil number was determined by Pearson’s correlation.

Acute myocardial infarction (AMI)

AMI was induced in adult wild-type female C57B6/J mice as previously described (*1*). Post-AMI peripheral blood cells, splenocytes, bone marrow and cardiac cells were isolated. Peripheral blood was obtained by cardiac puncture and combined with red blood cell (RBC) lysis solution (10 minutes at room temperature), peripheral blood mononuclear cells (PBMCs) were obtained by centrifugation 500 *g* for 10 minutes, followed by washing with phosphate buffered saline (PBS) (ThermoFisher Scientific) with 1 mM EDTA. A single cell suspension of splenocytes was produced by mashing intact spleen through a 70 µM cell strainer with the plunger of a 1 mL syringe into RBC lysis solution, RBCs were lysed for 10 minutes at room temperature and splenocytes centrifuged 500 *g* for 10 minutes. Bone marrow was isolated by flushing both femurs and tibias from each mouse with PBS using a 25G needle and a syringe. A single cell suspension of cardiac cells was produced by briefly mincing excised hearts with scissors and incubating with collagenase/dispase (Sigma-Aldrich) for 30 minutes at 37^o^C, followed by deactivation with the addition of 10% fetal bovine serum (FBS) (ThermoFisher, Scientific). Isolated single cell suspensions were utilised for flow cytometry analysis as detailed below.

miRNA-126 antagomiR

Adult wild-type female C57B6/J mice were pre-treated with miRNA-126 antagomiR (Qiagen) or scramble control at 0.5 mg / mouse by intraperitoneal injection 5 days and again 2 days prior to induction of AMI by LAD ligation.

Infarct size

Infarct size was calculated using 2,3,5-Triphenyl-2H-tetrazolium chloride (TTC) staining. Briefly, 24 hours post-AMI excised hearts were segmented into 1mm rings. Myocardial rings were incubated with TTC at 37^o^C and digitally imaged using a camera attached to a dissection microscope and by scanning PFA (4%) fixed myocardial rings with a digital scanner. Infarct size was calculated in Image J software by determining the total myocardial area, relative to the infarcted myocardium and expressed as a percentage. Both sides of each myocardial ring were imaged and a total of 14 measurements were averaged to obtain an infarct measurement per mouse. Analysis was conducted by operators who were blinded to study group allocations (scramble versus antagomiR).

Flow cytometry

5 x 10^5^ – 2 x 10^6^ cells were washed twice with buffer (PBS + 0.1 % BSA + 1 mM EDTA + 0.01% Sodium azide) by centrifugation 500 *g* at 4°C. Cells were Fc blocked with αCD16/CD32 (BD) or FcR-block Miltenyi Biotec, Ref. 130-092-575. Primary extracellular antibodies (**Table 1**) were added at indicated concentrations for 20 min at 4 °C in the dark and fixable viability Dye eFluor®780 (ThermoFisher) or 7-AAD viability stain (Invitrogen, Ref. 00-6993-50) was used to track for live cells. Labelled cells were washed and then fixed for 30 minutes using Cytofix (BD), washed again and resuspended in buffer before acquisition. CompBeads (BD) or single antibody staining controls were used to set up fluorophore compensation. Fluorescence minus one (FMO) controls were used to set gates for cell populations. Labelled cells were acquired using either an LSR II (BD), or Fortessa X20 (BD) flow cytometer using FACSDiva (BD). Data were analysed using Flowjo (Treestar, Inc.) software. After doublet and size exclusion, neutrophils were defined as live CD45^+^ CD11b^+^ Ly6G^+^ cells.

**Table 1 Antibodies**

| **Antibodies** | **Source** | **Identifier** |
| --- | --- | --- |
| Anti-CD45 BV650 (clone 30-F11) | BioLegend | Cat# 103151 RRID:AB_2565884 |
| Anti-CD11b BV510 (clone M1/70) | BioLegend | Cat# 101245 RRID:AB_2561390 |
| Anti-Ly6G Percp-Cy5.5 (clone 1A8) | BioLegend | Cat# 127615 RRID:AB_1877272 |
| Anti-Siglec F BV421 (clone E50-2440) | BD | Cat# 562681 |
| Anti-MHC II (I-A/I-E) Alexa Fluor 700 (clone M5/114.15.2) | eBioscience | Cat# 56-5321-82 RRID:AB_494009 |
| Anti-Ly6C PE-Cy7 (clone HK1.4) | eBioscience | Cat# 25-5932-82 |
| Fixable viability dye eFluor 780 | eBioscience | Cat# 65-0865-14 |
| Anti-CD45 AF647 (clone 30-F11) | BioLegend | Cat# 103124 |
| Anti- CD11b FITC (clone M1/70) | Biolegend | Cat# 101206 |
| Anti-Ly6G BV421 (clone 1A8) | Biolegend | Cat# 127636 |

Human and mouse endothelial cell extracellular vesicle generation, isolation and characterisation

Human umbilical vein endothelial cells (HUVECs) (Lonza, UK) and mouse endothelioma cell line sEND.1 were maintained in EGM-2 media supplemented with the BulletKit (Lonza, UK) in a humidified atmosphere with 5% CO_2_ or high glucose DMEM, with 10% FBS and 2 mM L-glutamine, respectively. All FBS supplementation was EV-depleted by ultracentrifugation for 16 hours at 120,000 *g* at 4^o^C. HUVECs were grown and maintained with a final concentration of 2% FBS and sEND.1 cells 10% FBS. Cells were treated with 10 ng / mL recombinant human or mouse tumour necrosis factor-α (TNF-α) (R&D Systems) at the described time points. EV were isolated as previously described (*1*). Briefly, cells were seeded into T75 cm^2^ or T175 cm^2^ tissue culture flasks at a density of 2 - 4 x 10^6^ / flask in 15mL EV-depleted medium and rested overnight. Subsequently, cell culture supernatants were removed and cell monolayers were washed with 15mL PBS and 15mL EV-depleted medium was added +/- (TNF-α) 10 ng / mL to each flask for 16 hours. Following cell culture stimulations supernatants were removed from cell culture flasks and centrifuged for 10 minutes at 1000 *g* to pellet cellular debris, the resulting supernatant was transferred to quick seal ultra-centrifugation tubes and centrifuged for 120,000 *g* for 2 hours at 4^o^C (Beckman Coulter, Optima MAX – XP Ultracentrifuge). EV pellets were resuspended in 100 µL PBS and were washed by combining isolated EV pellets with 13mL PBS and re-centrifuged 120,000 *g* for 1 hour. EV pellets were re-suspended in 100 µL PBS and utilised in studies as described.

VCAM-1 ELISA

Endothelial cell activation following stimulation with recombinant TNF-α (above) was confirmed in human and mouse cells using the human VCAM-1/CD106 DuoSet ELISA (R&D Systems) and mouse VCAM-1/CD106 DuoSet ELISA (R&D Systems) as per the manufactures instructions.

*in vivo* EC-EV studies

For *in vivo* EC-EV studies adult male B6SJLCD45.1 / C57BL10 / C57B6/J mice were used. mouse sEND.1 EC-EV were isolated as described above and labelled with *C*. *elegans* miR-39-3p transcript (Exiqon) using an exosomal transfection kit (Exo-Fect, System Bioscience) according to the manufacturer’s instructions as previously described (*1*). miR-39-3p labelled or unlabelled 1 x 10^9^ EVs in PBS were intravenously injected via the tail vein and blood/tissues collected at the described time points and utilised as described for quantification of neutrophil cell populations by flow cytometry, mRNA/miRNA RT-qPCR or for a protein chemokine array (details below). Briefly, for protein and RT-qPCR studies spleen tissue was macerated in lysis buffer (protein extraction in PBS with protease inhibitors (COmplete Tablets, Roche) or Qiazol for RNA) with a TissueLyser LT (Qiagen) at 50 oscillations twice for 1 minute each. Homogenized spleens were centrifuged at 4^o^C for 1 minute 8000 *g* and the supernatants collected for protein or RT-qPCR analysis.

Isolation of human neutrophils

Human peripheral blood neutrophils were isolated from STEMI and control NSTEMI patients at the time of presentation before percutaneous coronrary intervetion (PCI) and 1 month after AMI and total mRNA isolated for bulk RNA-sequencing.

Chemokine Array

The mouse proteome profiler mouse chemokine array kit (R&D Systems) was used to detect the expression of multiple chemokines in a single sample. 200 µg of splenocyte lysate was incubated with arrays as per the manufacturer’s instructions and imaged using the Bio-Rad ChemiDoc MP Imaging system. Image J software was used to determine the pixel density for the pairs of duplicate spots representing each chemokine. Data shown are chemokine array dot blot density values normalised to mean row value for each protein.

RT-qPCR mRNAs

Total RNA was isolated using miRNeasy Mini Kit (Qiagen) kit. cDNA was synthesised using QuantiTect Reverse Transcription RT-PCR kit (Qiagen). mRNA levels were determined by quantitative PCR (qPCR) using TaqMan probes (18s rRNA, *Gapdh, Cxcl12, Plxnb2, Il-1β, Cxcr2, Ccr2, Itag4, Cxcr4, Cxcl1, Il-6*) and reagents as per the manufacturer’s instructions. 18s rRNA was used as a housekeeper/normalization control and experimental data are shown as ΔΔCt values normalised to mean row ΔΔCt for each gene.

CRISPR-Cas9 base editing for VCAM-1

4 x 10^4^ sEND.1 cells were reverse transfected with 100 ng pX458-BE3 base editor plasmid (kind gift from Dr. Philip Hublitz, Radcliffe Department of Medicine, University of Oxford to Dr Joey Riepsaame, Sir William Dunn School of Pathology, University of Oxford.) harbouring a U6-sgRNA cassette targeting VCAM-1 (spacer: 5'-GATTCAATTCAGTGGCCCCC-3'), complexed with TransIT 2020 transfection reagent (Mirus) in OptiMEM as per the manufacturer's protocol. CRISPR-cas9 directed base editing (C>T) was utilised to introduce the Q122* premature STOP codon (CAA>TAA) in VCAM-1. Culture medium was replaced 16 hours after transfection and cells were allowed to recover for an additional 24 hours. Subsequently, targeted cells were enriched by fluorescently activated cell sorting (FACS) on (eGFP expression) and sorted as individual cells into a 96 well plate and cultured until they reached 70-100% confluence. Cells were expanded by replica-plating for cell banking and genomic DNA extraction and grown again to 70-100% confluence.

PCR-based genotyping of base-edited clones for VCAM-1

Genomic DNA was isolated by removal of the culture medium and lysing the cells for 2 hours at 55°C in lysis buffer (100 mM Tris pH 8.0, 5 mM EDTA, 200 mM NaCl, 0.2% SDS and 100 µg/mL proteinase K), followed by heating the samples for 10 min at 95°C. Genotyping was performed using Mismatch Amplification Mutation Assay (MAMA). In brief, the target region surrounding the intended point mutation for VCAM-1 was first PCR amplified using the primers VCAM-1_Fwd (5'-CGCCACAAACCATGACTAGC-3') and VCAM-1_Rev (5'-CGGCAAACAAGAGCTTTTCCA-3') to generate a 405 bp amplicon. PCR reactions comprised KAPA2G Fast Genotyping Mix (Sigma), isolated genomic DNA lysate, dH2O and genotyping primers. Thermal cycling was performed using the following conditions: 1 cycle 95°C for 3 minutes; 35 cycles 95°C for 10 seconds, 60°C for 10 seconds, 72°C for 10 seconds followed by final extension at 72°C for 1 minute. Upon exonuclease treatment of the PCR reaction, nested PCRs were performed on PCR amplicons (100 x dilution in dH_2_O) using the same PCR reagents, reverse primer and a forward primer specific to either the wild-type sequence ctttccccaaggatccagagattc (VCAM-1_Fwd_WT: 5'-ctttccccaaggatccagagatRC-3') or the point mutation ctttccccaaggatccagagattt (VCAM-1_Fwd_MUT: 5'-ctttccccaaggatccagagatMT-3'). Thermal cycling was performed using the following conditions: 1 cycle 95°C for 3 minutes; 20 cycles 95°C for 10 seconds, 60°C for 10 seconds, 72°C for 10 seconds; followed by final extension at 72°C for 1 minute. After PCR, samples were loaded onto a 0.8% agarose gel in TAE buffer and separated for 40 minutes at 100V. The resulting 247 base pair amplicons were cloned into pGEM-T vector (Promega) and DNA from 10 independent colonies was assessed by Sanger sequencing.

Nanoparticle tracking analysis of EVs

EV size and concentration profiles were determined by nanoparticle tracking analysis (NTA) using a Malvern Nanosight as previously described (*1*) or by the Particle Metrix NTA, Zetaview. Samples were diluted (1:1000) in 1 mL PBS and loaded into the sample chamber. Zetaview settings were: 11 positions, 2 cycles, medium video resolution. Pre-acquisition settings: sensitivity 80, fame 30, shutter speed 100. Post-acquisition settings min brightness 25, max size 1000, min size 5, trace length 15, nm/class 30, classes/decade64.

qPCR miRNAs

Isolated EC-EVs were combined with Qiazol reagent and EV-miRNAs isolated using the miRNeasy Mini Kit (Qiagen). cDNA was synthesised using the miScript II RT Kit (Qiagen) and miRNA primers hsa-126-3p, hsa-126-5p, mmu-126-3p, mmu-126-5p detected by qPCR using the miScript SYBR Green PCR Kit as per the manufacturer’s instructions. UniSp6 was used as a normalising control for miRNA RT-qPCR and spiked into the cDNA master mix as per the manufacturer’s instructions.

Transmission Electron Microscopy (TEM) and Cryo-TEM of EC-EVs

For negative staining TEM, grids (300 mesh Cu carbon film) were glow discharged for 20 seconds at 15 mA (Leica EM ACE 200). EC-EV samples were applied to the grid for 2 minutes, blotted, stained with 2% uranyl acetate for 20 seconds, blotted and allowed to air dry. Images were acquired on a 120kV Tecnai 12 TEM (ThermoFisher) equipped with a OneView digital camera (Gatan). For cryo-TEM imaging, grids (Quantifoil 200 mesh Cu 2/1 or lacey carbon grids) were prepared with a graphene oxide layer as previously described (*2*).  EC-EV samples were generated in T175cm^2^ flasks and isolated as described above. Isolated EC-EV were then applied for 2 minutes before being plunge frozen into ethane using a Vitrobot IV (ThermoFisher). Grids were imaged using SerialEM (*3*) on a Talos Arctica (operated at 200kV) equipped with a Falcon III camera (ThermoFisher).

Isolation and TEM of VCAM-1+ EVs

VCAM-1^+^ plasma EV were isolated as previously described (*1*). Briefly anti-human VCAM-1 antibodies (Clone # BBIG-V1, R&D Systems) were covalently conjugated to M-450 Tosylactivated Dynabeads as detailed in the manufacturer’s instructions. 500 µL of platelet poor plasma was combined with 13mL PBS and ultra-centrifuged for 2 hours at 120,000 *g* and EV re-suspended in 100 µL PBS. Anti-VCAM-1 or IgG magnetic beads of iron oxide (MPIOs) were incubated with isolated plasma EV at room temperature for 1 hour under continual rotation. After incubation, MPIOs were pelleted for 5 minutes using a Dynal Magnet (Invitrogen), supernatants were discarded and MPIOs washed 3 times in 1 mL of PBS. VCAM-1^+^ EV-MPIOs were fixed with 1.6% glutaraldehyde in PBS for 1 hour at 4^o^C and then washed with PBS embedded in 4% low melting point agarose. The agarose was cut into small cubes (1-2 mm^3^) and processed for TEM (*4*). Sections were collected onto 200 mesh Cu grids and imaged using a Gatan OneView camera with a FEI Tecnai 12 TEM at 120kV.

Western blotting

EC-EV protein markers were confirmed as previously described (*1*). Supernatants from 4 x T175 cm^2^ flasks of endothelial cells were pooled for 1 sample/lane on described gels/membranes. Cell pellets, EV-depleted supernatants and cell culture media that was not incubated with cells but contained EV-depleted FBS was used as controls. EC-EV were lysed using 1x RIPA buffer (Cell Signalling Technology) with protease (C0mplete Tablets, Roche) and phosphatase inhibitors (PhosSTOP, Roche) and needle sonicated. 10-40 µg of protein was loaded per sample after combining with NuPage LDS sample buffer (4x) agent (Invitrogen) with and without 10x reducing agent (Invitrogen). EC-EV protein was loaded onto a 4-12% bis-tris gradient gel (NuPAGE 4-12% Bis-Tris Protein Gel; 1.5 mm (ThermoFisher Scientific) under non-reducing or reducing conditions (NuPAGE™ Sample Reducing Agent (10X)). Separated samples were transferred to nitrocellulose membranes and blocked for non-specific binding in 5 % milk in PBS-tween for 1 hour. Membranes were incubated with primary antibodies overnight: ALIX (ab117600, Abcam) (1/2000 dilution), TSG101 (ab83, Abcam) (1/2500 dilution), CD9 (EXOAB-KIT-1, System Biosciences) (1/2000 dilution) anti-VCAM-1 (EPR5047, Abcam) (1/2000), anti-ATP5A (15H4C4, Abcam) (1/5000), anti-histone H3 (D1H2/4499P, Cell signalling) (1/2000) and anti-eNOS (BD, Biosciences) (1/2000) in 5 % milk in PBS-tween. Membranes were washed 3 times and incubated with secondary-horse radish peroxidase (HRP) conjugated antibody for 1 hour. Membranes were washed again before incubating with enhance chemiluminescence substrate (Pierce ECL, ThermoFisher Scientific) for imaging (Bio-Rad ChemiDoc MP Imaging system).

*In silico* bioinformatics for miRNA-126-mRNA target prediction and neutrophil Gene Ontology pathway analysis

miRNA-126-predicted mRNA targets were obtained from TargetScanHuman, TargetScan (*5*), miRWalk (*6*), miRDB (*7*) and compared to Gene Ontology (GO) (*8, 9*) terms deposited for neutrophil biology as described. Gene lists and GO term lists were compared by Fisher’s exact test to determine significant overlap using all protein-coding genes or all GO terms

Isolation of human neutrophils

Peripheral venous blood collected in to EDTA tubes was diluted 1:1 with PBS and neutrophils were negatively enriched using the EasySep direct human neutrophil isolation kit as per the manufacturers instructions. Isolated neutrophils were with lysed with RLT (Qiagen) and stored at -80^o^C.

Neutrophil RNA isolation and RNA-sequencing

RNA was isolated using RNeasy Mini Kit (Qiagen), according to manufacturer’s instructions. The amount of total RNA was quantified using the Qubit Fluorometric Quantitation system (Life Technologies) and the RNA integrity number (RIN) was determined using the Experion Auto-mated Electrophoresis System (Bio-Rad). RNA-seq libraries were prepared with the TruSeq Stranded mRNA Library Prep kit (Illumina) using both Sciclone and Zephyr liquid handling robotics (PerkinElmer). Library concentrations were quantified with the Qubit Fluorometric Quantitation system (Life Technologies) and the size distribution was assessed using the Experion Automated Electrophoresis System (Bio-Rad). For sequencing, samples were diluted and pooled in equimolar amounts and sequenced on Illumina HiSeq 3000/4000 instruments in 50-bp-single-read configuration. Base calls provided by the Illumina Real-Time Analysis (RTA) software were subsequently converted into BAM format (Illumina2bam) before de-multiplexing (BamIndexDecoder) into individual, sample-specific BAM files via Illumina2bam tools (1.17.3. https://github.com/wtsi-npg/illumina2bam). Libraries were prepared and sequenced by the Biomedical Sequencing Facility (BSF) at CeMM.

### RNA-sequencing differential gene expression

Transcript abundance quantification was carried out from raw RNA-seq reads by Salmon v0.14.0 (*10*) with GC bias correction and mapping validation options, using the Ensembl human cDNA annotation release 96 (*11*). Analysis of differential gene expression was carried out using DESeq2 v1.24.0 (*12*) with a multi-factor experimental design comparing paired presentation vs. follow-up data for each patient and controlling for group (STEMI or NSTEMI). Briefly, pre-filtering was performed to remove cDNAs with fewer than 10 total reads across all patients. Differential expression analysis was then carried out with the default parameters. Shrinkage of log fold changes for visualisation in MA plots was carried out using the *apeglm* R package (*13*).

### Gene Set Enrichment Analysis

The fGSEA v1.10.0 R package (*14*) was used to detect enriched gene sets within the STEMI presentation vs. follow-up comparison. The hallmark gene sets v6.2 from the Molecular Signatures Database (*15*) were tested for enrichment. Genes were ranked according to the p-value for differential expression obtained from DESeq2, with negative fold changes assigned a negative value. fGSEA was run with 20,000 permutations and restricted to gene sets with between 10–500 genes.

**Single Cell RNA-sequencing**

Single cell RNA-sequencing was undertaken as previously described (*16, 17*). Briefly, viable Ter119-B220-NK1.1-CD11b+ cells were sorted from blood or the heart of male C57BL6/J mice following: control (N = 3), sham surgery following 1 day (N = 3) and AMI following 1 day (N = 5), using a FACS Aria III (BD Biosciences) with a 100µm nozzle. The proportion of neutrophils (Ly6G+) and non-neutrophils (Ly6G-) were adjusted to approximately 1:1 proportions. Samples were labelled with Hashtag antibodies 1 to 7 (Biolegend TotalSeq-A) for multiplexed sing cell RNA-sequencing analysis (*18*) and loaded in the 10x Genomics Chromium for sing cell RNA-sequencing library preparation. Libraries were generated with the 10x Genomics Chromium Single Cell 3’ Reagents Kit v3 Chemistry and sequenced on a Novaseq 6000 system (Illumina).

**GO term enrichment in single cell neutrophil clusters following AMI**

Differentially expressed genes from mouse neutrophil single cell clusters following AMI were assessed for enrichment against the mouse genome of GO terms (separately for biological process, molecular function, and cellular component and Reactome pathways).

**Comparison of human neutrophil and mouse neutrophil transcriptomes following AMI**

Differentially expressed genes from 3 mouse single cell clusters following AMI were individually compared to human differentially expressed genes in STEMI presentation vs. follow-up by Fisher exact test to determine significant overlap between the datasets. A background of 25000 total genes was used for the calculations.

**Comparison of enriched GO terms in human and mouse neutrophils following AMI**

Significantly enriched GO terms from mouse peripheral single cell clusters following AMI were individually compared to human differentially expressed genes in STEMI presentation vs. follow-up by Fisher exact test to determine significant overlap between the datasets. Backgrounds of 29112 total biological processes, 11118 molecular functions, 4181 cellular components or 2362 Reactome pathways were used for the calculations, respectively.

**Enrichment of miRNA-126-mRNA targets within neutrophil transcriptomes following AMI**

The miRNA-126-predicted mRNA target lists (human, mouse and human-mouse overlap) obtained as previously described were compared to human differentially expressed genes in STEMI presentation vs. follow-up by Fisher’s exact test. The same analysis was performed comparing miRNA-126-predicted mRNA targets to differentially expressed genes from mouse neutrophil single cell clusters following AMI. A background of 25000 genes was used in calculations.

**References**

1.	N. Akbar, J. E. Digby, T. J. Cahill, A. N. Tavare, A. L. Corbin, S. Saluja, S. Dawkins, L. Edgar, N. Rawlings, K. Ziberna, E. McNeill, S. Oxford Acute Myocardial Infarction, E. Johnson, A. A. Aljabali, R. A. Dragovic, M. Rohling, T. G. Belgard, I. A. Udalova, D. R. Greaves, K. M. Channon, P. R. Riley, D. C. Anthony, R. P. Choudhury, Endothelium-derived extracellular vesicles promote splenic monocyte mobilization in myocardial infarction. *JCI Insight* **2**, (2017).

2.	M. Thomas G., B. Andreas, W. P. F. Anthony, S. Sjors H. W., *Graphene Oxide Grid Preparation*. (2016).

3.	D. N. Mastronarde, Automated electron microscope tomography using robust prediction of specimen movements. *Journal of Structural Biology* **152**, 36-51 (2005).

4.	T. Riffelmacher, A. Clarke, F. C. Richter, A. Stranks, S. Pandey, S. Danielli, P. Hublitz, Z. Yu, E. Johnson, T. Schwerd, J. McCullagh, H. Uhlig, S. E. W. Jacobsen, A. K. Simon, Autophagy-Dependent Generation of Free Fatty Acids Is Critical for Normal Neutrophil Differentiation. *Immunity* **47**, 466-480 e465 (2017).

5.	V. Agarwal, G. W. Bell, J. W. Nam, D. P. Bartel, Predicting effective microRNA target sites in mammalian mRNAs. *Elife* **4**, (2015).

6.	C. Sticht, C. De La Torre, A. Parveen, N. Gretz, miRWalk: An online resource for prediction of microRNA binding sites. *PloS one* **13**, e0206239 (2018).

7.	W. Liu, X. Wang, Prediction of functional microRNA targets by integrative modeling of microRNA binding and target expression data. *Genome Biol* **20**, 18 (2019).

8.	The Gene Ontology Consortium, The Gene Ontology Resource: 20 years and still GOing strong. *Nucleic acids research* **47**, D330-D338 (2018).

9.	M. Ashburner, C. A. Ball, J. A. Blake, D. Botstein, H. Butler, J. M. Cherry, A. P. Davis, K. Dolinski, S. S. Dwight, J. T. Eppig, M. A. Harris, D. P. Hill, L. Issel-Tarver, A. Kasarskis, S. Lewis, J. C. Matese, J. E. Richardson, M. Ringwald, G. M. Rubin, G. Sherlock, Gene ontology: tool for the unification of biology. The Gene Ontology Consortium. *Nature genetics* **25**, 25-29 (2000).

10.	R. Patro, G. Duggal, M. I. Love, R. A. Irizarry, C. Kingsford, Salmon provides fast and bias-aware quantification of transcript expression. *Nature methods* **14**, 417-419 (2017).

11.	D. R. Zerbino, P. Achuthan, W. Akanni, M R. Amode, D. Barrell, J. Bhai, K. Billis, C. Cummins, A. Gall, C. G. Girón, L. Gil, L. Gordon, L. Haggerty, E. Haskell, T. Hourlier, O. G. Izuogu, S. H. Janacek, T. Juettemann, J. K. To, M. R. Laird, I. Lavidas, Z. Liu, J. E. Loveland, T. Maurel, W. McLaren, B. Moore, J. Mudge, D. N. Murphy, V. Newman, M. Nuhn, D. Ogeh, C. K. Ong, A. Parker, M. Patricio, H. S. Riat, H. Schuilenburg, D. Sheppard, H. Sparrow, K. Taylor, A. Thormann, A. Vullo, B. Walts, A. Zadissa, A. Frankish, S. E. Hunt, M. Kostadima, N. Langridge, F. J. Martin, M. Muffato, E. Perry, M. Ruffier, D. M. Staines, S. J. Trevanion, B. L. Aken, F. Cunningham, A. Yates, P. Flicek, Ensembl 2018. *Nucleic acids research* **46**, D754-D761 (2017).

12.	M. I. Love, W. Huber, S. Anders, Moderated estimation of fold change and dispersion for RNA-seq data with DESeq2. *Genome Biology* **15**, 550 (2014).

13.	A. Zhu, J. G. Ibrahim, M. I. Love, Heavy-tailed prior distributions for sequence count data: removing the noise and preserving large differences. *Bioinformatics* **35**, 2084-2092 (2018).

14.	G. Korotkevich, V. Sukhov, A. Sergushichev, Fast gene set enrichment analysis. *bioRxiv*, 060012 (2019).

15.	A. Liberzon, C. Birger, H. Thorvaldsdóttir, M. Ghandi, Jill P. Mesirov, P. Tamayo, The Molecular Signatures Database Hallmark Gene Set Collection. *Cell Systems* **1**, 417-425 (2015).

16.	E. Vafadarnejad, G. Rizzo, L. Krampert, P. Arampatzi, V. A. Nugroho, D. Schulz, M. Roesch, P. Alayrac, J. Vilar, J.-S. Silvestre, A. Zernecke, A.-E. Saliba, C. Cochain, Time-resolved single-cell transcriptomics uncovers dynamics of cardiac neutrophil diversity in murine myocardial infarction. *bioRxiv*, 738005 (2019).

17.	G. Rizzo, E. Vafadarnejad, P. Arampatzi, J.-S. Silvestre, A. Zernecke, A.-E. Saliba, C. Cochain, Single-cell transcriptomic profiling maps monocyte/macrophage transitions after myocardial infarction in mice. *bioRxiv*, 2020.2004.2014.040451 (2020).

18.	M. Stoeckius, S. Zheng, B. Houck-Loomis, S. Hao, B. Z. Yeung, W. M. Mauck, 3rd, P. Smibert, R. Satija, Cell Hashing with barcoded antibodies enables multiplexing and doublet detection for single cell genomics. *Genome Biol* **19**, 224 (2018).
